# The Genetic and Proteomic Determinants of Pediatric Stature Development and their link to adult height and Type 2 Diabetes

**DOI:** 10.1101/2025.11.10.25339894

**Authors:** N Fragoso-Bargas, AE Lupu, I Campillo-Pereda, Y Huang, J Sundfjord, R Karimi, T Lind, JC Holm, LA Holm, A Havdahl, OA Andreassen, S O’Rahilly, P Sole-Navais, B Jacobsson, E Bratland, G Davey-Smith, PB Juliusson, KK Ong, T Hansen, PR Njølstad, M Vaudel, S Johansson

## Abstract

Stature is a key indicator of pediatric health, yet the molecular and metabolic architecture governing childhood growth remains incompletely defined. We conducted genome-wide association analyses of stature from birth to childhood (574,580 measurements; 72,704 Norwegian children) and integrated UK Biobank data (age 10 and adulthood). We uncovered life-stage shifts in the genetics of stature, identifying novel childhood-specific effects including variants near *GLP1R* with BMI-independent effects, and reveal three temporal clusters - infancy and childhood clusters enriched for endocrine and metabolic pathways, and a lifetime growth cluster enriched for skeletal biology. Proteome analysis revealed that prepubertal height is associated with widespread changes in the circulation, identifying 106 height-associated proteins and linking IGF1, IGF2, and IGFBP3 levels to childhood-height polygenic scores. Mendelian randomization showed that reduced fetal growth and taller prepubertal height increase type 2 diabetes risk, independent of adult stature. These findings uncover developmental stage-specific genetic and proteomic influences on child-growth and links to adult metabolic health.

## INTRODUCTION

Physical growth is a universal indicator of an infant or child’s overall health and nutritional status and is closely monitored in pediatrics (1,2). Stature, assessed through supine length or standing height, is a simple and accessible measurement that, alongside other clinical parameters, can help identify children with underlying chronic diseases or undernutrition, as it allows for evaluation of growth patterns using standardized growth curves (2). The Infancy-Childhood-Puberty (ICP) growth model defined by Karlberg (3) divides post-natal growth into three phases; Infancy: from birth up to approximately 3 years; Childhood: starting during the first year up to 11 years; and Puberty: representing the additional growth during puberty. Nutrition predominantly influences infancy, thyroid hormone thyroxine is active during infancy and childhood periods, sex steroid hormones during puberty, while growth hormone is active across the three phases (3). These phases therefore reflect different growth biology; however, their genetic underpinnings remain to be systematically characterized.

Despite the scientific and clinical importance of stature development in early life, the genetics of normal-height has mainly been studied in adults and adolescents. The largest genome-wide association study (GWAS) of adult height to date involved 5.4 million individuals (4). The study found 12,111 independent SNPs associated with adult height, explaining approximately 40% of the variance in individuals of European ancestry, representing nearly the entire contribution of common variant influences on final height (4). Thus, adult height is now considered the most comprehensively genetically characterized human trait. The Early Growth Genetics (EGG) consortium identified 10 loci associated with height during early puberty (n = 18,737), assessed at mean age 10 in girls and 12 in boys, as well as for height change and peak growth velocity between 10-12 years and adulthood (5). More recently, a study (n = 56,000) identified 26 genome-wide signals involved in pubertal growth during early adolescence (6).

In contrast, the genetic architecture of stature during earlier life stages remains uncharted. Two GWASs of length at birth identified one and ten loci in 28,459 and 154,000 babies, respectively (8). How genetic variation shapes height development in infancy and early childhood has not been systemically assessed. Twin studies have shown that heritability of height follows an evolving pattern, increasing from low levels at birth (10-15%) to around 50% at age 1 year and continues to increase throughout early childhood (9,10).

Previous studies have suggested contrasting positive and negative links between stature and type 2 diabetes (T2D), depending on timing of growth. Rapid height gain after 7 years of age is associated with higher risk of glucose intolerance and T2D in later life (11,12) and a GWAS of pubertal height indicated a positive genetic correlation with fasting insulin levels, HOMA-IR, T2D, and other indicators of impaired metabolic health (6). Conversely, both longer length at birth and taller adult height have been associated with lower risk for T2D (13,14). A better understanding of the processes underlying the complex relationship between height development and glucose homeostasis related traits is needed.

In regards to the plasma proteome, childhood height remains poorly characterized. Adult height has been examined alongside multiple traits in the plasma Proteome Atlas, covering approximately 3,000 proteins in up to 50,000 individuals from the UK Biobank (15). In contrast, only one study has assessed proteomics in relation to childhood height (16). This study was conducted in a Nepalese population of undernourished children aged 6-8 years, including around 500 individuals and 982 proteins. The study identified key components of the IGF axis such as IGF1, IGFBP3, and IGFALS (16). However, it is unclear how representative these findings are for well-nourished children and other populations. Hence, the relationship between the plasma proteome, healthy childhood growth, and their potential association with childhood genetics remains unknown.

In this study, we performed GWAS in up to 72,704 children from the longitudinal Norwegian Mother, Father and Child (MoBa) study. By using 574,580 length/height measurements from birth to age 8 years, and integrating the results with UK biobank height data at age 10 and adulthood, we provide new insights of the shared and timing-specific genetic architecture acting on stature across the life-course and the complex relationship between early growth and later metabolic health. We compared how child-based and adult-based polygenic scores associate with childhood height development and to clinically relevant height biomarkers using three additional population-based child-cohorts (ALSPAC - age 7- 9 years, Holbækk age 5-9 years, and OTIS (6, 12 and 18 months old)) and we investigated the association between 1,365 circulating proteins and height in children aged 4 to 9 years.

## RESULTS

### The genetic trajectories of stature, from birth to adulthood

GWASs were performed for length/height measurements across 12 time points from birth (n = 72,704) to age 8 years (n = 27,924) (Figure 1a and Supplementary table 1) and compared with self-recalled comparative height at age 10 (HAC10, n = 332,021) and adult height (n = 458,303) in the UK BioBank (13). The SNP-based heritability of length/height increased markedly from birth (h2 = 0.16, se = 0.01) to age one year (h2 = 0.31, se = 0.02), and then more gradually to age 8 years (h2 = 0.41, se = 0.03), when it was similar to that of adult height (h2 = 0.38, se = 0.02) (Figure 1b and Supplementary table 2). The between-age genetic correlations were generally strong (Figure 1c and Supplementary table 3) but decreased with increasing time between measurements (birth vs adult: rg = 0.52; 8y vs adult: rg = 0.73), indicating both shared and distinct genetic architectures across time. The genetic architecture could grossly be described as three overlapping phases with particularly strong intra-pairwise correlations: birth to 3-months, 3 to 16-months, and 16-months to 8 years.

**Figure 1:**
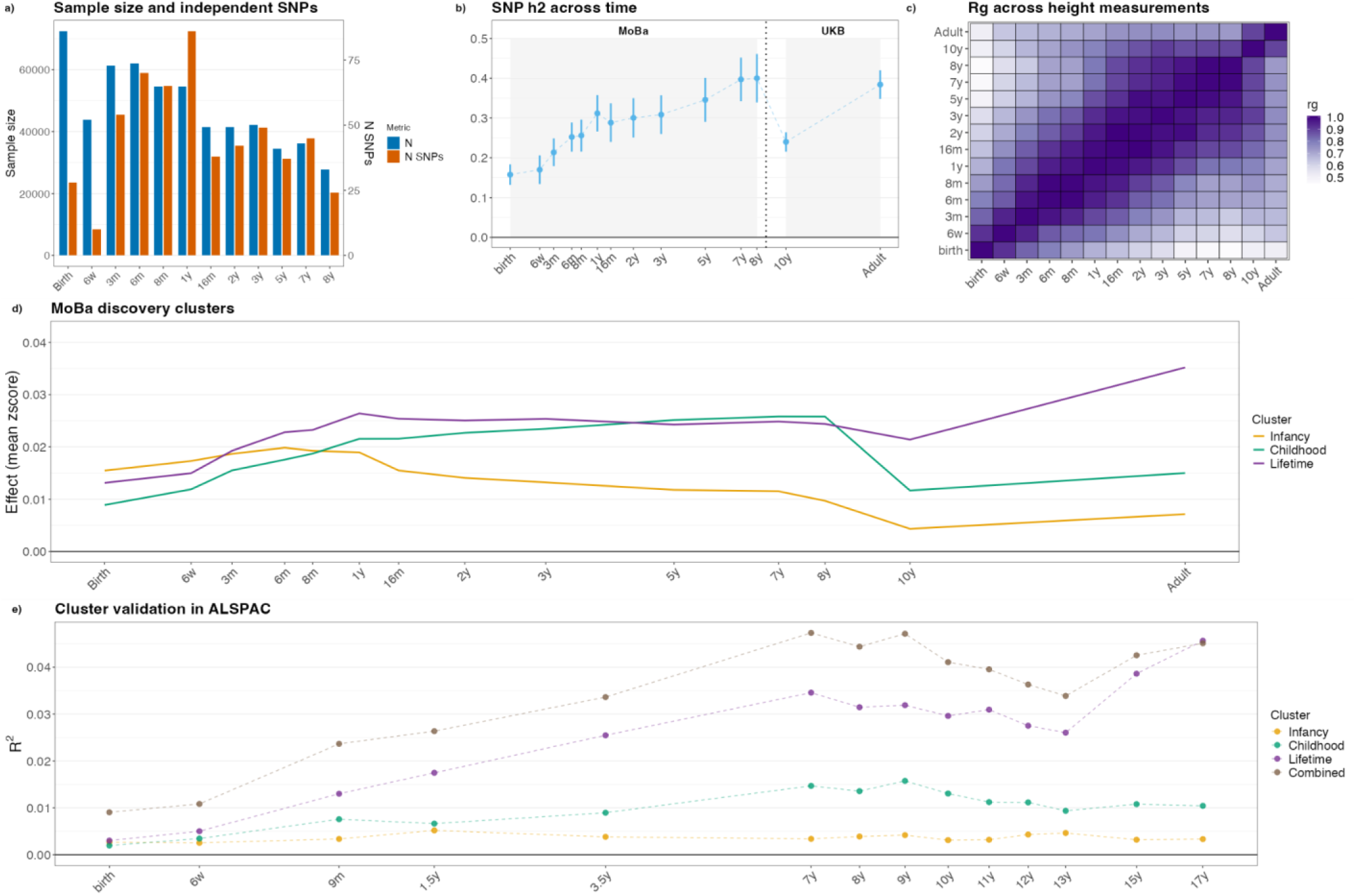
GWAS of childhood height across 12 timepoints and cluster analysis. a) Sample size at each time-point, and numbers of independent SNPs per time-point. b) Increasing SNP-based heritability of length/height across MoBa time-points, UKBB comparative height at 10-years and adult height. c) Genetic correlations across all time points. d) Averaged effect sizes (Z-scores) of hierarchically clustered lead SNPs at each time-point. e) Variance explained in height in ALSPAC by lead SNPs within the *Infancy*, *Childhood* or *Lifetime* clusters, and all 179 SNPs combined.

We identified between 24 and 86 conditionally independent signals (using GCTA-COJO) at each timepoint in the MoBa sample (Figure 1a, Supplementary table 4). Pairwise colocalization (COLOC) analysis yielded 179 independent signals across all time points (Figure 1a and Supplementary table 5). Of these 179 signals, 98 and 129 reached genome-wide significance also for self-recalled comparative height at age 10 (HAC10) and measured adult height in the UK Biobank, respectively (Supplementary figure 1 and Supplementary table 6). An additional 28 and 9 SNPs showed directionally-concordant effects at a Bonferroni corrected threshold (p < 0.05/179) on HAC10 and adult height, respectively. Conversely, 22 of the 179 MoBa signals showed weaker associations with HAC10 or adult height (Supplementary table 6). Of these, 19 lead SNPs were not in linkage disequilibrium (LD, r2<0.6) with SNPs associated with birth length (8) or adult height in the largest published meta-analyses (4), and none was associated with height in the GWAS Catalog. However, the nearest genes to some SNPs have been linked to traits such as adult height, bone density, and T2D (Supplementary table 7). Interestingly, two novel signals (rs72857945 and rs579786) were near *DNAH8/GLP1R*, the latter being relevant for appetite and adiposity regulation. Notably both signals have stronger effects during infancy. Their individual trajectories across MoBa time-points are shown in Supplementary figure 2.

We used hierarchical clustering of effect estimates across the 12 MoBa timepoints, HAC10, and adult height to explore the between-age dynamics of the 179 MoBa length/height signals. This revealed 3 clusters of signals with distinct effect trajectories across ages (Figure 1d and Supplementary table 5): 1) *Infancy* (n = 60 signals): predominant effects from birth to 1-year; 2) *Childhood* (n = 71): stronger effects from 1- to 8-years than at 10-years and on adult height, and 3) *Lifetime* (n = 48): consistent effects across ages 1- to 10-years and larger effect on adult height. Of the 19 novel signals for human height, 16 were assigned to the *Infancy* cluster and 3 to the *Childhood* cluster (Supplementary figure 2). Of the 16 novel signals available in ALSPAC, 12 showed directionally-concordant effects on length at the 9-month time-point, which best represents the effects of signals in the *Infancy* cluster (Sign test p = 0.03) (Supplementary table 7). We constructed polygenic scores (PGS) for each of the 3 clusters and a combined PGS including all 179 variants, and estimated the variance explained in height from birth to age 17 years in ALSPAC (Figure 1e and Supplementary table 8). The variance explained trajectories in ALSPAC aligned well with the cluster profiles in MoBa (Figure 1e), although the *Infancy* PRS explained little variance in height in ALSPAC.

### The genetic architecture of stature in early-life is largely similar between sexes

We performed sex stratified GWASs in girls (n = 35,560 to 13,749 from birth to 8-years) and boys (n = 37,144 to 14,175), along with a joint model including a SNP-by-sex interaction term. SNP-based heritability increased with age in both sexes, with only slightly higher estimates in girls (Supplementary figure 3a and Supplementary table 9). Genetic correlations between girls and boys were close to one from birth to age 2-years, and declined modestly thereafter, suggesting largely shared genetic architecture with minor divergence before puberty (Supplementary figure 3b and Supplementary table 10).

At their peak associated time point, none of the 179 loci identified in the combined analysis showed evidence of a sex-specific effect (Supplementary Table 11), and effect sizes were highly concordant with those in sex-combined models (Supplementary figure 3c), with no sex-interaction effects identified (Supplementary Table 11). Across the genome, no SNP exhibited a genome-wide significant sex interaction. The sex stratified GWAS identified 48 signals in girls and 37 in boys, including a few not identified in the sex-combined model (9 in boys and 15 in girls). Only rs17213081, identified in girls, reached the stricter threshold for sex stratified analyses (p at birth = 3.6×10^⁻9^; Supplementary table 12), but its isolated genomic location, borderline significance (p at birth = 3.2×10^⁻9^), and lack of LD with SNPs associated with relevant traits render this signal inconclusive. Overall, we find no robust evidence for sex differences in the genetic architecture of length/height from birth to 8-years.

### The performance of a PGS based on adult height variants increases over time

We constructed a polygenic score (PGS) for adult height using GWAS summary statistics from over 1.5 million individuals of European ancestry, based on the meta-analysis by Yengo et al. (4), including approximately 10,000 independent lead SNPs. We then evaluated its predictive value across childhood within MoBa (birth to 8-years, Figure 2a and Supplementary table 13) and ALSPAC (birth to 17-years, Figure 2b and Supplementary table 13). The adult height PGS performed similarly in MoBa and ALSPAC and across both sexes with increasing performance from birth (variance explained: 4%) to age 8-years (18-22%) (Figure 2a and 2b). From ages 9/10- to 13/15-years in ALSPAC, corresponding to the period of pubertal maturation in both sexes, there was a notable drop in performance (Figure 2b). This reduction was more pronounced and occurred approximately 2 years later in boys than in girls, consistent with the later timing of the pubertal growth spurt in boys. Overall, the adult PGS explained a greater proportion of the variance than the childhood-based PGS even at the infancy and early childhood time-points.

**Figure 2:**
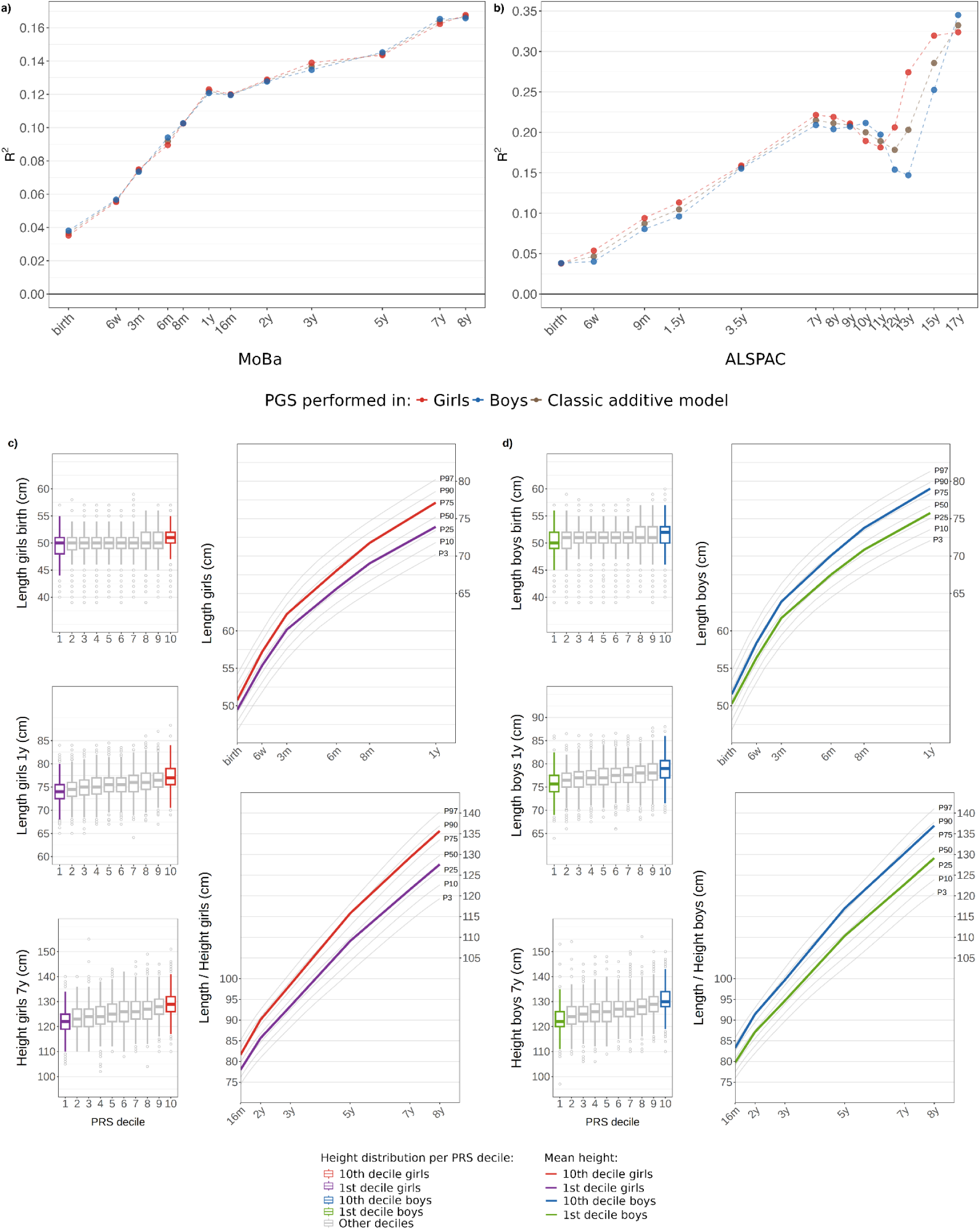
Variance explained (R²) by adult height SNPs in childhood and adolescent height, and longitudinal height trajectories across PGS Deciles in MoBa: a) Variance in infancy and childhood length/height in MoBa explained by an adulthood height PGS. b) Similar analysis in ALSPAC. c) Height distribution by decile in girls with first decile in purple and tenth decile in red, and stature development chart including the mean for first (purple) and tenth decile (red) at birth, 1 year old and 7 years old. d) Height distribution by decile in boys with first decile in green and tenth decile in blue, and stature development charts including the mean for first (green) and tenth decile (blue) at birth, 1 year old and 7 years old. See Supplementary figure 4 for trajectories of individuals drawn at random to illustrate the consistent height growth pattern in 1st and 10th decile.

Over time, growth-curves diverged increasingly between children with low (PGS < 10th percentile) and high (PGS > 90th percentile) PGS for adult height (Figure 2c and 2d). At age 1-year, girls in the top adult height PGS decile were on average 3.1 cm (1.21 SD) taller than those in the bottom decile. By age 7-years, the difference increased to 7.5 cm (1.39 SD) corresponding to the 89th and 28th percentiles in high vs low adult height PGS groups according to the standard Norwegian growth charts. Boys showed a similar pattern, with a 3.2 cm difference at age 1-year (1.21 SD) and a 7.7 cm difference at age 7-years (1.42 SD). A set of individual growth charts are presented in Supplementary figure 4.

We next evaluated whether the adult height PGS improves height prediction beyond the classical clinical predictors: mother’s height, father’s height, and their combination (Figure 3 and Supplementary table 14). The variance explained by the adult height PGS increased over time; it was a better predictor than single parental height, but worse than the combined parental heights (Figure 3 and Supplementary table 14). The addition of the PGS to combined parental heights consistently improved the total variance explained, with the largest improvement observed at age 7-years, increasing from 21% to 26%.

**Figure 3:**
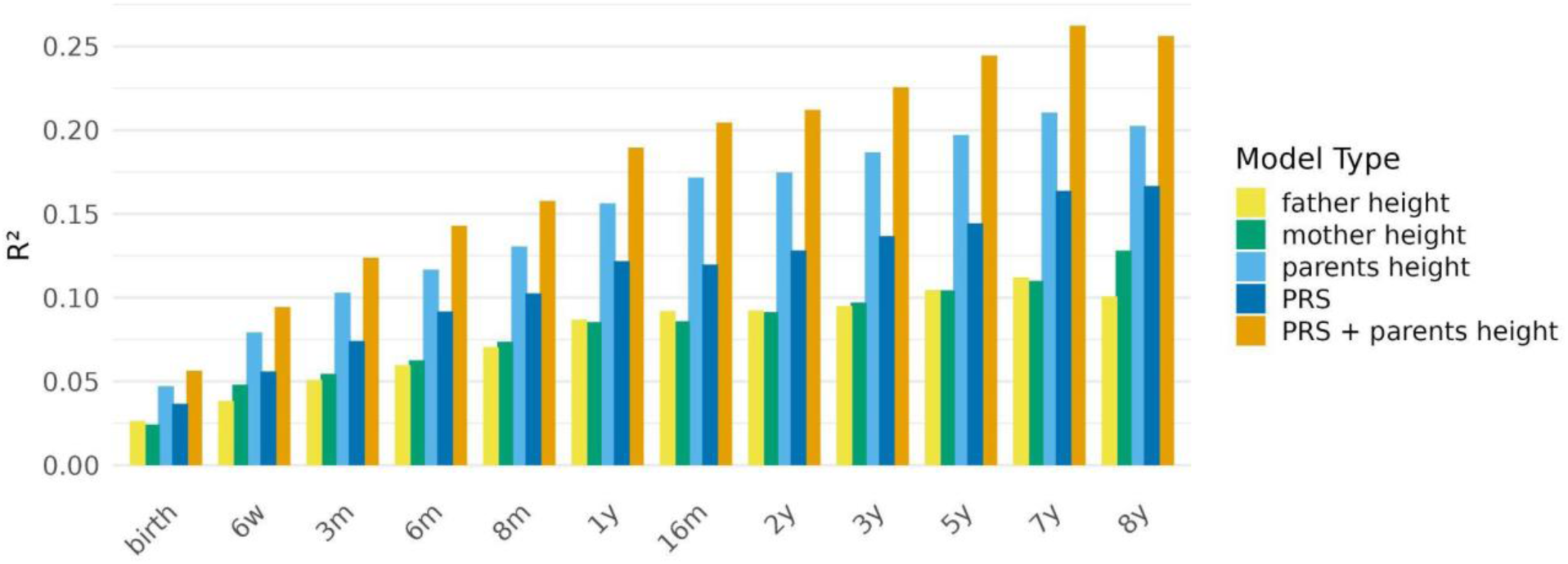
Variance explained (R²) on height across all MoBa time points using combinations of different predictors. Models constructed: father’s height (yellow): Stature ∼Father stature+sex; mother’s height (green); Stature ∼ Mother stature + sex; parents’ heights (light blue): Stature ∼ Mother stature + Father stature + sex; Adult-height PGS (strong blue): Stature ∼ PGS + sex; Adult-height PGS + parents’ heights (orange): Stature ∼ PGS + Mother stature + Father stature + sex.

### Early life stature development links to distinct mechanisms and diseases

The timing of effects across the three SNP clusters broadly aligned with Karlberg’s three stages of growth, prompting us to investigate whether these clusters reflect distinct underlying biological mechanisms. Mapping the 179 lead SNPs to their nearest genes allowed us to conduct gene expression enrichment analyses that revealed significant enrichment (FDR < 0.05) for growth disorders, bone diseases, and endocrine disorders such as insulin resistance and type 2 diabetes (Figure 4a and Supplementary Table 15). While all three clusters showed enrichment for growth related disorders, only the *Infancy* and *Childhood* clusters were enriched for metabolic diseases and endocrine disorders, and only the *Lifetime* cluster was enriched for bone diseases (Figure 4a and Table 1, Supplementary table 16 and 17).

**Figure 4:**
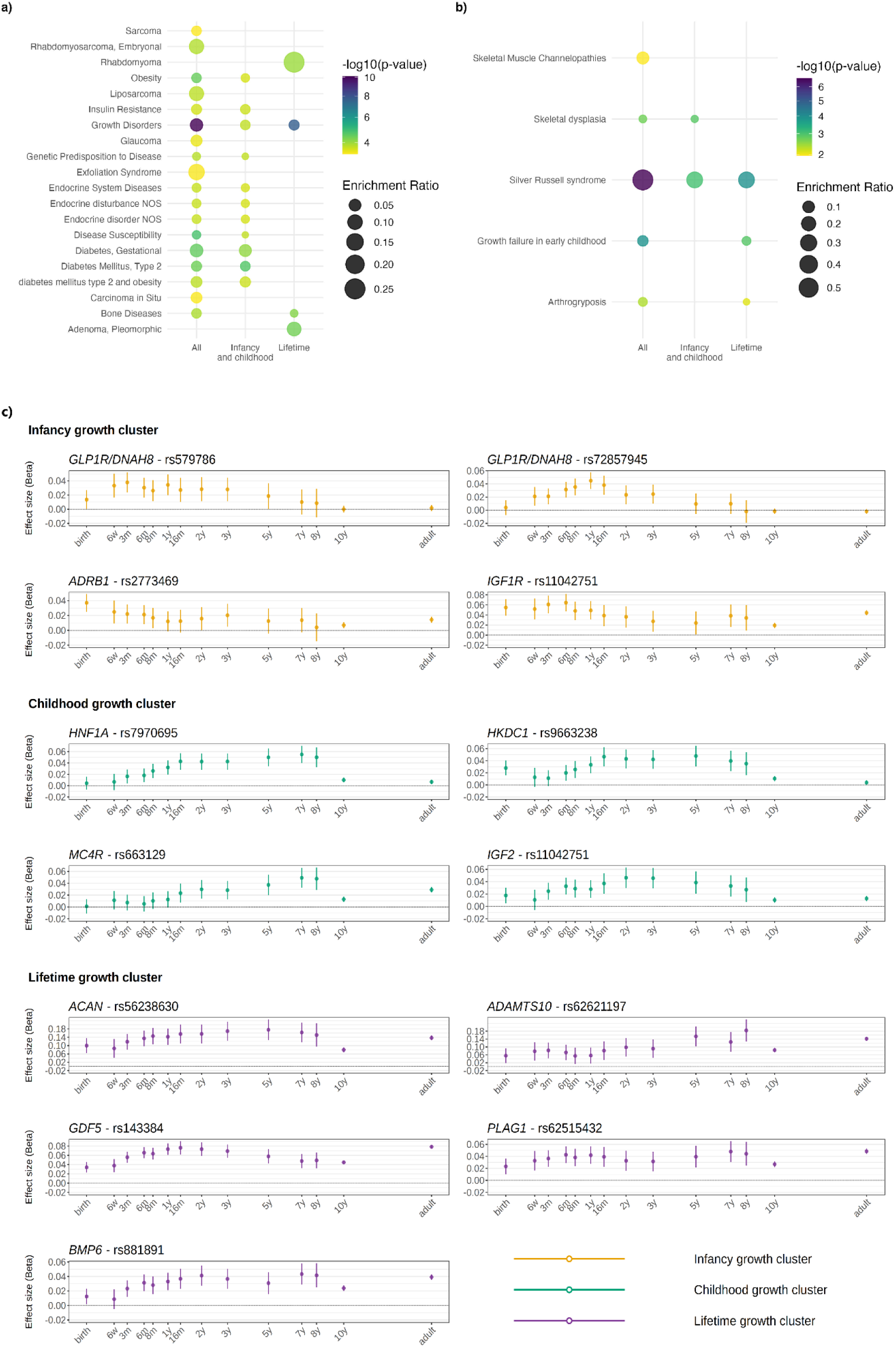
Enrichment analyses and effect trajectories of variants mapped to selected biologically relevant genes: GLAD4U a) and b) UK rare variant panel enrichment analyses. All displayed biological terms reached a 5% FDR threshold in at least one analysis. e) Individual SNP effect size (95% CI) trajectories for selected driver-genes from enriched pathways.

**Table 1:**
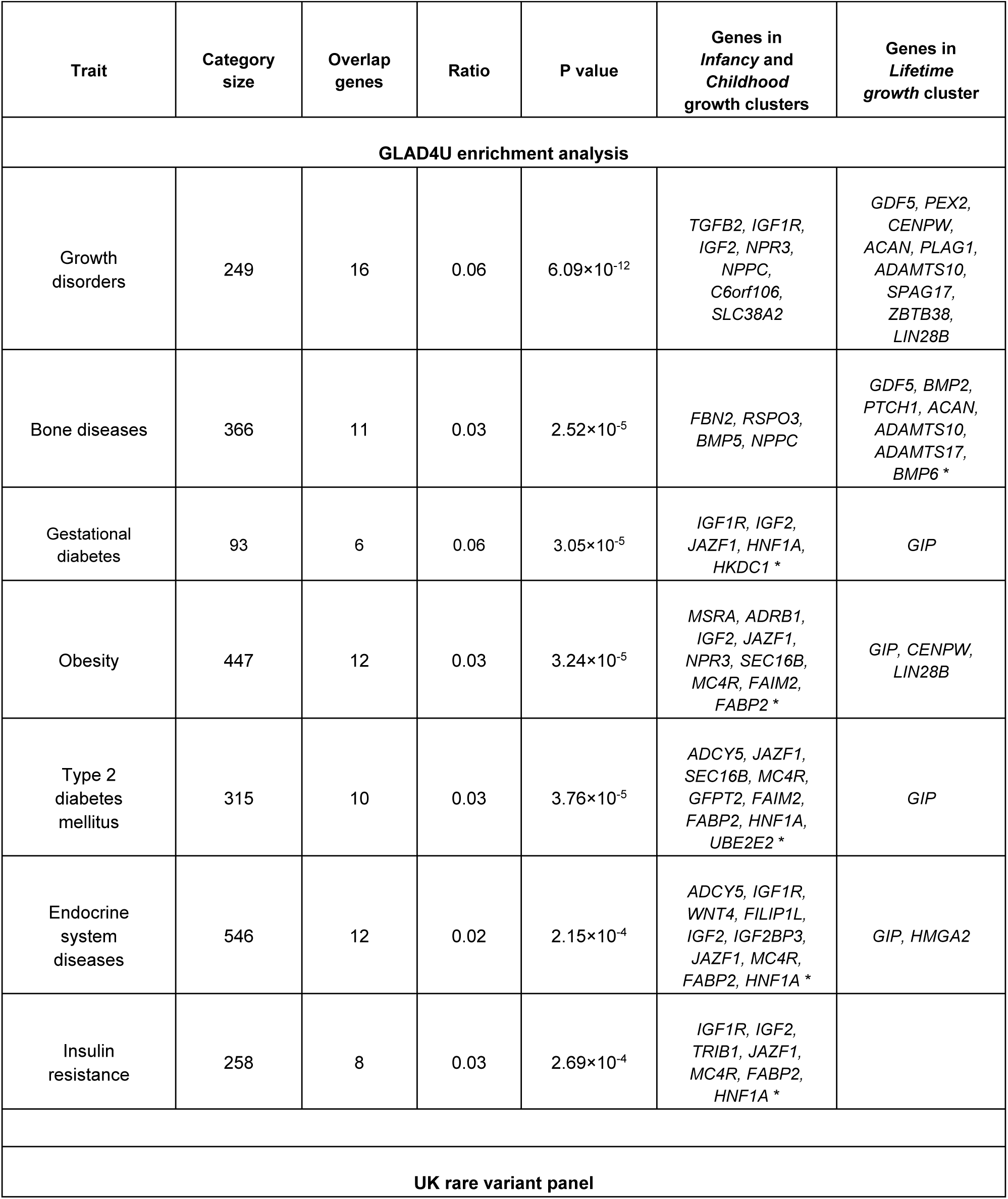
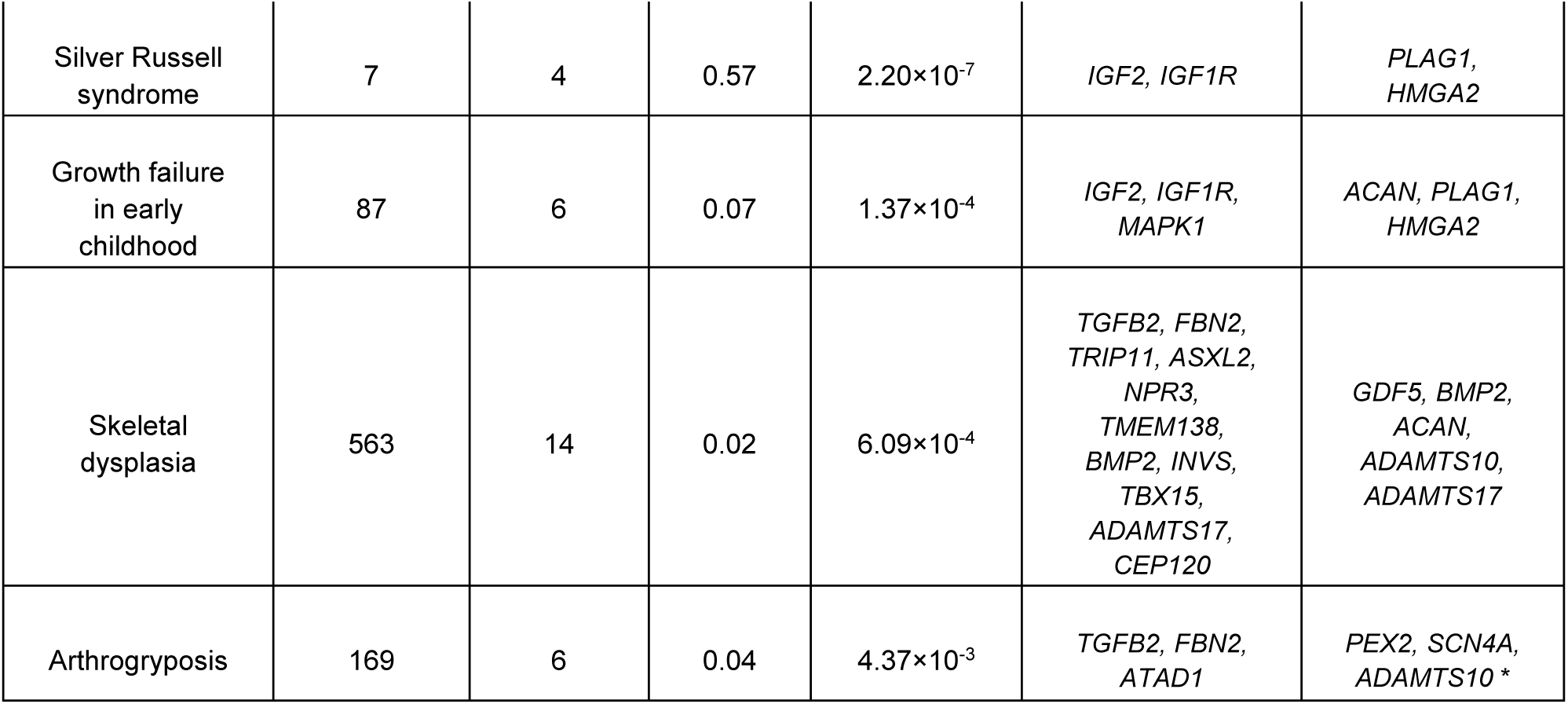
Overview of lead traits from the GLAD4U and UK rare variant panel enrichment analyses and the driver-genes listed according to cluster classification (more extensive details are shown in supplementary tables 9-14). The number of genes overlapping with the trait-category, and their corresponding enrichment ratio and p value are combined for all three clusters. Individual genes are listed according to their cluster category and cluster classes, and categories marked with * are statistically significantly overrepresented only in the respective cluster.

A more focused enrichment analysis for Mendelian growth-related disorders from the UK rare variant panels revealed enrichment for “Silver Russell syndrome”, “Skeletal dysplasia”, “Growth failure in early childhood”, and “Arthrogryposis” (Figure 4b and Supplementary tables 18-20). Key enriched categories and the driver-genes from each cluster are displayed in Table 1, and the complete growth trajectories of selected driver genes are shown in Figure 4c. Tissue enrichment analysis in FUMA using the Gene to Function analysis on Genotype-Tissue Expression (GTEx) tissue expression datasets on all the SNPs (n = 179) showed upregulation in the tibial artery, tibial nerve, adipose visceral omentum and subcutaneous adipose tissue (Supplementary table 21 and Supplementary figure 5).

### Cross-trait correlations and Mendelian Randomization reveal links to metabolic traits

We chose length/height measurements at birth, 1-year and 7-years in MoBa, as well as comparative height at age 10 (HAC10) and adult measurements in UK Biobank, to represent distinct developmental phases. For these time points, we calculated genetic correlations with anthropometric, adiposity, pubertal, and glucose-related traits (Figure 5a and Supplementary Table 22). Birth weight correlated strongly with birth length (rg = 0.71, se = 0.04), but also showed positive genetic correlations with length/height at later time-points, indicating a substantial shared genetic contribution between height and fetal weight development. Length, already at age 1-year, displayed positive correlations with trunk and body fat percentage and T2D, and negative correlations with age at puberty (menarche/first facial hair). Height at 7-years (prepuberty) showed positive correlations with adult BMI and body fat, recalled comparative adiposity/body size at age 10, fasting insulin, and negative correlations with age at puberty. This indicates that genetic mechanisms that promote taller stature before puberty may also contribute to increased later risk for impaired glucose homeostasis and metabolic disease.

**Figure 5:**
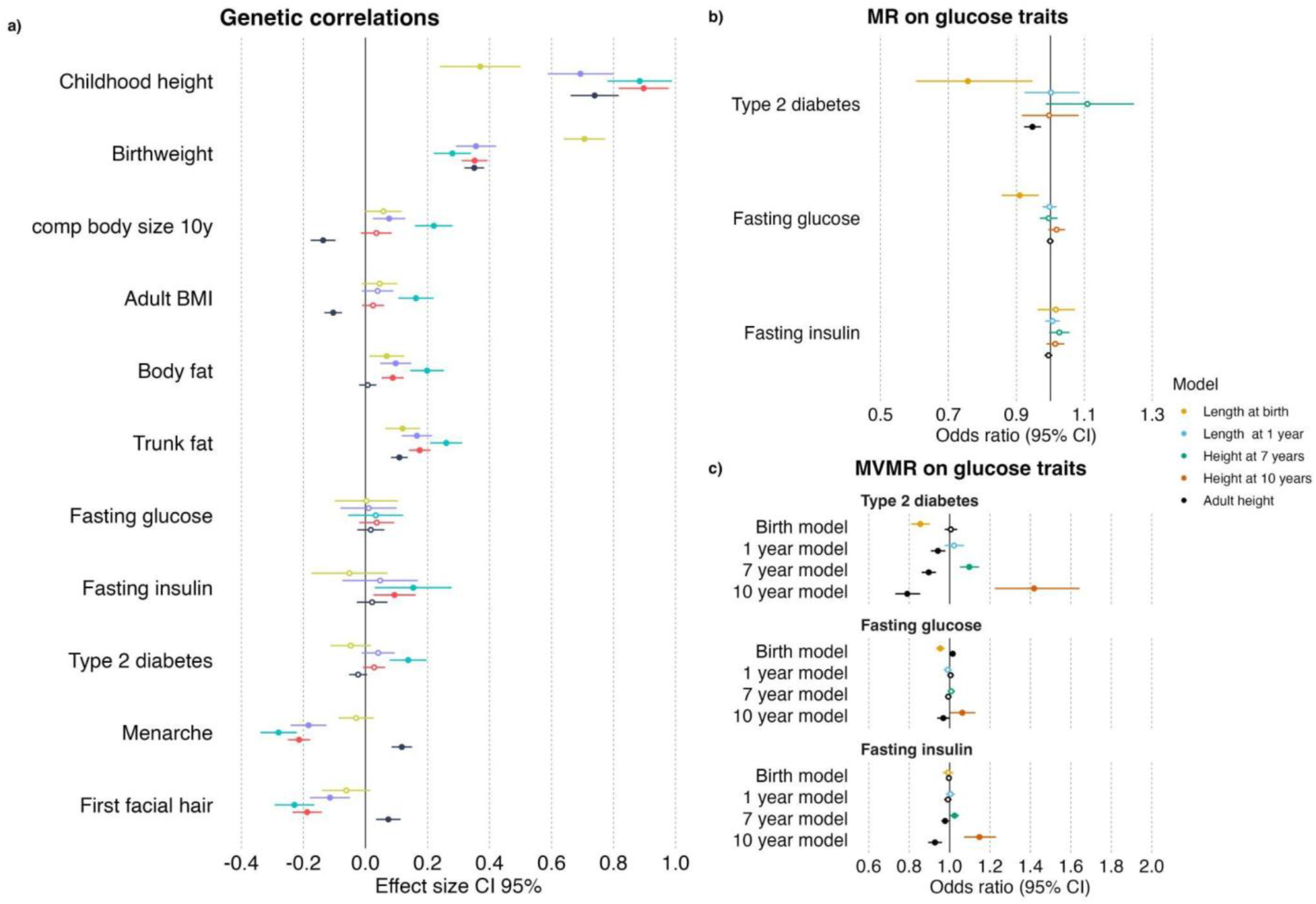
Genetic correlations with other traits and Mendelian randomization analyses between height across time and glucose homeostasis related traits. a) Genetic correlations between height at representative time-points (birth, 1, 7 and 10 years, adult) and other traits. b) Inverse variance weighted univariable Mendelian randomization analysis. c) inverse variance weighted multivariable Mendelian randomization analysis of childhood height at representative time-points adjusted for adult height. In the MVMR analysis, the child effect indicates the estimated causal impact of childhood height on the outcome after accounting for adult height. Conversely, the adult height effect indicates the causal impact of adult height after accounting for the childhood height. In all Forest plots, dots indicate effect size estimates. Filled dots indicate statistically significant associations. Abbreviations: MR: Mendelian randomization. MVMR: Multivariable Mendelian randomization.

The positive genetic correlation between height at 7-years and T2D contrasts with the established inverse phenotypic and genetic associations between adult height and T2D risk (12). Indeed, univariable Mendelian randomization analyses indicated that both genetically predicted shorter birth length and shorter adult height are associated with higher T2D risk (Figure 5b and Supplementary table 23). Multivariable Mendelian randomization, including both birth length and adult height in the same model, showed a predominant effect of birth length on T2D (MVMR OR = 0.85, CI: 0.81-0.90) and no direct effect of adult height (Figure 5c and Supplementary table 24). Similar results were obtained for fasting glucose. These results suggest that the observed association between taller adult height and lower risk for T2D is more likely attributable to mechanisms that promote fetal growth. In contrast, similar multivariable Mendelian randomization analyses accounting for adult height indicated that taller height at 7- and 10-years conferred higher risk for T2D and higher fasting insulin levels. Both multivariate weighted median and EGGER agreed with the multivariable inverse variance weighted (IVW) analysis, supporting the robustness of this analysis (Supplementary table 24).

### Genetic pre-disposition to height associates with protein levels of IGF1, IGF2 and IGFBP3 in children

To examine the mechanisms by which genetic predisposition impacts childhood height, we performed a targeted analysis of classical growth-related proteins and metabolites measured at ages 5 to 9 years in ALSPAC (Supplementary table 25). The biomarkers tested were IGF1, IGF2, IGFBP3, Leptin, fasting glucose, SHBG, GHBP, and fasting insulin (n = 423 to 4798). We tested association with the childhood height PGS (n= 179 SNPs) and the adult height PGS (n= ∼11k SNPs). At a 5% FDR threshold, IGF1, IGF2, and IGFBP3 were positively associated with the childhood PGS, whereas no biomarker was associated with the adult PGS, albeit with weaker trends in the same directions (Figure 6a and Supplementary table 25). Sensitivity analyses using PGSs based on the three childhood height clusters showed similar results (Supplementary table 26). We then replicated these findings in healthy children aged 4 to 9-years from the Danish HOLBAEK Study (n = 239), observing highly similar results for IGF1 and IGF2, again with slightly stronger effects for the childhood height PGS compared to the adult height PGS (Figure 6b, Supplementary table 27). We also tested associations with infancy levels of IGF1 and IGFBP3 in the Swedish OTIS study (n= ∼200, at ages 5, 12 and 16-months) and found the childhood height PGS was associated with IGFBP3 levels at both 5 and 16 months (Figure 6b, Supplementary table 27).

**Figure 6:**
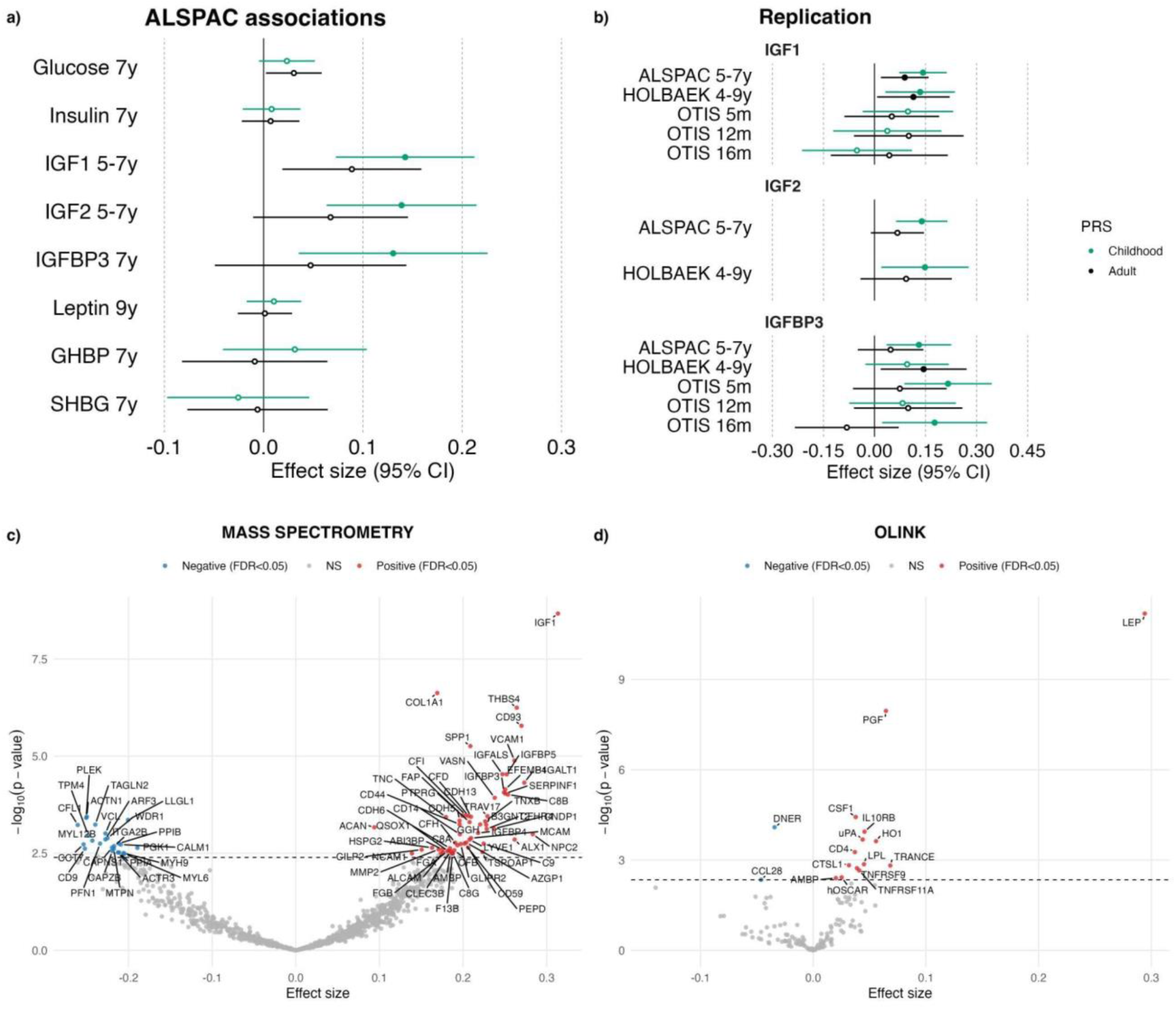
Associations of child height and adult height polygenic risk scores with growth-and metabolism-related phenotypes and height-associated circulating proteins. a) Associations between child height and adult height PGSs and relevant growth and metabolic phenotypes in the ALSPAC cohort. b) Replication of IGF-related protein associations in the HOLBAEK and OTIS cohorts. c) proteome-wide associations with measured childhood height in HOLBAEK using proteins quantified by mass spectrometry and d) the OLINK panels, respectively.

We next performed a targeted analysis of 22 anthropometric and metabolite traits in HOLBAEK (n = up to 634 children, Supplementary table 28). Height and weight were positively associated (FDR<0.05) with both PGSs, while waist-to-height ratio (WHR), BMI and LDL cholesterol were negatively associated only with the adult height PGS (supplementary table 28). Cluster-specific sensitivity analyses (Supplementary Table 29) confirmed childhood height PGS associations (p < 0.05) with height and weight, but not with adiposity-related traits such as BMI and WHR.

Lastly, we extended the childhood and adult height PGSs association analyses to the broader proteome measured by mass spectrometry (MS) (1,216 proteins, n= ∼240 children) and two Olink targeted panels including 149 inflammation- and cardiovascular-related proteins (n= ∼630 children) in the HOLBAEK Study (Supplementary tables 30 and 31). Only 5 proteins, ACAN, PTGD5, VWA5B2, STXBP5, CLEC3B (all from MS), showed an association (at FDR < 0.05), and all were positively associated with the adult height PRS.

### Proteomic analysis identifies proteins associated with measured height at 4 to 9-years

We performed a hypothesis-free screen in the population-based cohort of the HOLBAEK Study, testing measured child height against 1,365 circulating proteins. Child height was associated (FDR < 0.05) with 82 proteins measured by MS (Figure 6c and Supplementary Table 32) and 16 proteins measured with Olink (Figure 6d and Supplementary Table 33).

Among the top associated proteins measured by MS were Insulin-like growth factor-1 (IGF1) and its binding partners IGFBP3, IGFBP5 and IGFALS. Child height also showed positive correlations with proteins related to bone formation pathways, such as COL1A1, SPP1, ACAN, and THBS4. Among inflammation- and cardiovascular-related proteins measured in Olink panels, the top associated proteins were LEP and PGF (Placenta Growth Factor). Significant protein-height associations were further adjusted for WHR in sensitivity analyses, showing that effect directions were preserved in both MS and OLINK datasets (Supplementary Tables 34 and 35). Notably, most protein-height associations did not attenuate after conditioning for WHR, including the strong persisting positive association with LEP (p= 8.22 ×10^-11^).

Protein-associations were not enriched among any biological pathways at FDR < 0.05. However, with a more lenient FDR threshold of 10%, several biologically relevant pathways showed enrichment, including, osteochondrodysplasias, metabolic bone diseases, and bone density. Within the metabolic bone diseases and bone density categories, implicated effector proteins included IGF1, IGFBP3, IGFALS, COL1A1, and ACAN. All biological terms that reached p<0.05 are shown in Supplementary table 36.

## DISCUSSION

In this study, we performed GWASs of 574,580 child height measurements from 72,704 children from the Norwegian longitudinal MoBa birth cohort and extended these analyses with proteomics data from three population-based child cohorts. As the largest genetic and proteomics study of childhood height to date, our results address a knowledge-gap in our understanding of this fundamental and universal health indicator.

Our results extend previous large-scale genetic studies in adulthood by characterizing the gradually evolving genetic control of growth from relatively low levels at birth and early infancy, followed by a rapid increase in heritability from around 3 months to 1 year, and then a more gradual rise up to 8 years of age, when SNP-based heritability reaches levels comparable to adulthood. Although a considerable component of the genetic architecture is shared between adult and child stature, our data also suggests that some of the genetic landscape changes along with childhood phases, likely reflecting more life-long and more temporally important mechanisms respectively as has been suggested by the Karlberg model (3).

Further support for this concept comes from the three life-course effect trajectory-clusters identified for the 179 identified lead SNPs. Our first and second clusters align with the classical Karlberg growth phases, *Infancy* (birth to 3-years) and *Childhood* (1 to 11-years), with effects that attenuate throughout puberty (3,17). The third, *Lifetime* cluster reflects genetic influences with lasting effects on height and likely represents loci responsible for the considerable genetic overlap seen across time. The smaller size of this cluster is probably a consequence of the gradual decline in participation rate with age in MoBa, favouring discovery of early-life associated loci. Considerably larger child-samples are needed to formally test this, but it is reasonable to assume that a large fraction of adult-discovered loci will fall in this group. The third Karlberg growth phase, pubertal growth, could not be directly mapped as MoBa discovery GWASs were performed only up to age 8-years, but this likely explains the notable decline in PRS performance between ages 10 and 15 seen in the ALSPAC cohort.

While the *Lifetime* cluster presented enrichment of skeletal related traits and disorders, as expected for stature, the *Infancy* and *Childhood* clusters share enrichment for pathways related to glucose homeostasis and endocrine functions. Within the *Infancy* cluster are signals near obvious biological candidates such as *IGF2*, as well as signals near *IGF1R* and *ADRB1* with well known associations with diabetes and obesity (18,19). In addition, novel insights include two independent signals (rs72857945 and rs579786) at the *DNAH8/GLP1R* locus that influence height only in infancy and early childhood. Here, the most likely effector gene is *GLP1R*, important for glycemic control and appetite regulation, and a drug target for diabetes and obesity (20). Notably, these two lead SNPs for height are not in high LD (r^2^ < 0.5) with the three nearby childhood BMI-signals that we reported previously (21) that show a similar transient effect restricted to early childhood adiposity. Furthermore, we noted that our height signal rs72857945 is highly correlated with the recently discovered semaglutide weight-loss associated rs10305420 (r² = 0.70), and with rs9357296 (r² = 0.98), linked to nausea symptoms during semaglutide treatment (22). Notably, the alleles corresponding to taller infancy height are associated with less vomiting and lower weight-loss during treatment. This highlights that our approach is able to identify pharmacogenomically relevant SNPs and illustrates how variants associated with early-life traits may influence specific phenotypes in adulthood.

*GLP1R* may influence early childhood growth via its roles in insulin signaling and central energy regulation. We speculate that the lead height SNPs rs579786 and rs72857945 may express their effects via promotion of insulin secretion through *GLP1R* agonists, and thereby induce growth hormone secretion (23). However, *GLP1R* may also play a more multifaceted role in early growth, by also modulating bone resorption via calcitonin stimulation in thyroid C cells (24). This highlights its broader physiological relevance, suggesting that *GLP1R* linked variants may influence growth through both metabolic and skeletal pathways.

In the *Childhood* cluster, we identify a signal near *MC4R,* a key component of the leptin-melanocortin pathway with known impacts on obesity (25) and childhood BMI (21,26). Another interesting gene within this cluster is *HKDC1,* involved in glucose metabolism pathways and linked to gestational diabetes (27–29). *HKDC1* may also impact growth through its role in cellular homeostasis to prevent cellular senescence (30). Lastly, in the *Lifetime* cluster, we highlight *BMP6* and *ACAN,* both well known height related loci that have been shown to affect bone and cartilage growth (31–33), consistent with their strong effects during childhood and lifelong.

We did not observe major differences in the genetic architecture of stature between girls and boys in early childhood, suggesting that their genetic underpinnings, like the observed traits, may start diverging with the onset of puberty (34,35). Similarly, the PGS are consistent in girls and boys up to 7-years after which there is a clear shift in performance, suggesting potential genetic mechanisms influencing height through puberty timing, more so in boys where it differs from girls by approximately 2 years.

The adult PGS explained approximately 20% of height variance at ages 8 to 9-years in both ALSPAC and MoBa, increasing to 33% at age 17 in ALSPAC. These findings indicate that although adult genetic influences are already substantial in late childhood, the genetic architecture of height continues to develop and diverge from that of adulthood throughout adolescence. The notable drop in performance of both the adult height and *Lifetime* cluster PGSs during puberty and subsequent better performance in adolescence in ALSPAC highlights the role of a distinct set of genetic factors influencing height during puberty, which current MoBa data cannot capture.

Overall, our childhood height PGSs were less powerful than the adult height PGS due to lower statistical power of our discovery sample compared to the more than 5 million participants for adult height. Notably, the adult height PGS impacts length/height already from birth. This reflects the influence of lifelong SNP effects that begin to manifest early in development, consistent with the considerable genetic correlation (rg) between height at different ages and the rise of the *Lifetime* cluster effects from age 1-year. However, growth in childhood is also shaped by age-specific genetic and environmental factors. Some adult SNPs may be misweighted when applied to early life stages, and certain SNPs appear to exert effects only during childhood, as evidenced by the 19 SNPs in the *Infancy* cluster that have no detectable influence on adult height. These findings highlight the dynamic nature of height genetics across the life-course and the importance of considering developmental context when interpreting PGS performance. Still, the classical model based on both parents’ height performed better at predicting childhood height compared to the pure genetic model, highlighting an influence of factors that are not captured by common variation. Both predictors showed increasing performance across early life, peaking around ages 7 to 8-years, when parental heights explained ∼20% and the adult height PGS ∼15% of the variance. In combination, the explained variance increased noticeably to ∼26%, highlighting the potential of genetic information to complement traditional growth monitoring and enhance early clinical decision-making.

Mendelian randomization analyses revealed insights into the life-course relationship of height/length with T2D, fasting glucose, and insulin levels. The protective association between taller adult height and glycemic traits disappears when accounting for birth length, suggesting that the causal relationship between stature and glycemic risk originates in early life. Indeed, we found a robust protective link between greater birth length and T2D or fasting glucose in both univariable and multivariable MR. This aligns with the previously reported negative relationship between birth weight and T2D risk (36), which can be explained by genetic determinants of reduced insulin secretion causing both lower fetal growth (weight and length) and higher later T2D risk (36,37). Adding to the complexity, our results also indicate that a taller prepubertal childhood height relative to final adult height increases T2D risk. Although these relationships may seem contradictory, previous literature supports that rapid height gain from 7 to 13-years may pose higher risk for disordered glucose homeostasis (12), while individuals with early-onset puberty tend to have relatively shorter adult heights (32). Moreover, this aligns with the observed genetic correlations, particularly the negative relationships between pubertal traits and length already from age 1-year onwards.

As previously noted, the adult-based height-PGS explains considerably more of childhood height variance than the childhood-PGS that was based on our much smaller MoBa-cohort. However, our protein and metabolite association analyses in ALSPAC showed that IGF1, IGF2, and IGFBP3, factors central in both growth (39,40) and insulin resistance related pathways (41–43), showed slightly stronger associations with the child height PRS than with the adult height PRS, despite the latter containing far more SNPs. These findings were replicated in HOLBAEK (IGF1 and IGF2) and OTIS (IGFBP3). Lastly, in the agnostic, hypothesis-free proteome association analysis of measured height, we identified IGF1 and IGFBP3, along with other IGF-axis proteins such as IGFBP5 and IGFALS, reinforcing the key role of the IGF-axis in statural growth and highlighting shared biological mechanisms linking growth and metabolic health.

Among the measured height-protein associations, we also note several proteins involved in bone formation. These include COL1A1, which is responsible for making part of type I collagen (44), the most abundant protein in bone, and SPP1 (Osteopontin) a key secreted phosphoprotein and bone matrix component important for bone remodelling (45), ACAN which is associated with adult height (4), and THBS4 involved in cartilage integrity and angiogenesis (45,46). The OLINK analysis identified two proteins better known for other key roles: LEPTIN and PGF. LEPTIN, well known for its role in central energy regulation, has increased levels among children with obesity and our data provides some support for the argument that leptin might be a putative mediator of the increased prepubertal growth seen in children with obesity. Our data shows that leptin is associated with height even after controlling for WHR and studies have shown that leptin has both direct anabolic effects on osteoblasts and chondrocytes, and indirect influences on bone via the hypothalamus and sympathetic nervous system (47). The strong association between taller stature and higher levels of PGF (Placenta Growth Factor), better known for angiogenesis and vasculogenesis, suggests that the importance of PGF goes beyond stimulating osteoclast differentiation during fracture healing (48). Other notable height associated proteins include several key components of the monocyte-macrophage-osteoclast formation and activity pathway i.e. TRANCE (known as RANKL), TNFRFS11A (RANK), CSF1, OSCAR and IL10RB (49,50).

These findings collectively underscore the importance of ossification for child growth, whereby newly formed bone is remodelled to grow in length, thickness, and strength.

A key strength of this study is its use of data from one of the largest and most comprehensive longitudinal birth cohorts in the world - the MoBa study. We included a total of 574,580 height measurements, performed by trained health nurses, from birth to age 8-years, making this the largest GWAS for early life height. Data from parents allowed us to evaluate the performance of predictive models that combined PGS with parental heights. Moreover, the availability of multiple measurement time-points allowed us to explore the differing likely causal relationships over time with T2D and related traits. The use of replication data (ALSPAC) further validated the robustness of our findings. Furthermore, we provide evidence on how specific proteins associate with both polygenic liability to childhood height and measured childhood height, relationships that have not been previously characterized in healthy children. However, limitations remain: although we identified 179 signals at conventional GWAS significance, the sample size is considerably smaller than in adult height GWASs and considerably larger childhood sample sizes are needed to achieve power comparable to the adult height PGS. Moreover, our sample was exclusively of European ancestry, and future research should include diverse populations to widen the generalizability of the results.

To conclude, our results shed new light on the genetics of statural growth, and suggest mechanistic links between early life height and later glycaemic related traits. Moreover, the time-dependent dynamics observed in genetic correlations, MR analyses and IGF protein associations, highlight the importance of conducting GWAS at developmental stages, in addition to adulthood, to better understand the relationships between complex traits and disease risks.

## METHODS

### Study populations

#### MoBA

The Norwegian Mother, Father and Child Cohort Study (MoBa) is a pregnancy-based ongoing cohort study consisting of approximately 114,500 children, 95,200 mothers and 75,000 fathers, enrolled from 50 hospitals across Norway from 1999 to 2008 (51,52). In 41% of the pregnancies invited, the pregnant women consented to participation. The establishment of MoBa and initial data collection was based on a license from the Norwegian Data Protection Agency and approval from The Regional Committees for Medical and Health Research Ethics. The MoBa cohort is currently regulated by the Norwegian Health Registry Act. The current study was approved by The Regional Committees for Medical and Health Research Ethics (#2012/67).

#### ALSPAC

The ALSPAC (Avon Longitudinal Study of Parents and Children) cohort has been extensively described elsewhere (53,54), In brief, it initially recruited pregnant women residing in Avon, UK, with expected delivery dates between April 1, 1991, and December 31, 1992. A total of 14,541 pregnancies were enrolled, resulting in 14,676 fetuses, of which 13,988 children were alive at one year of age. When the children reached age seven, an additional 906 pregnancies were recruited, increasing the sample size for analyses from age seven onwards to 15,477 pregnancies. These additional pregnancies resulted in 15,658 fetuses, with 14,901 children alive at one year. Initially, 14,203 unique mothers participated in the study, with 630 more women joining at the 7-year follow-up, bringing the total to 14,833 mothers. Partners of the mothers were also invited to participate; 12,113 partners have been in contact with the study, and 3,807 are currently enrolled. Up to ∼6,500 children have both genotype and height data available. The study website contains details of all the data that is available through a fully searchable data dictionary and variable search tool (www.bristol.ac.uk/alspac/researchers/our-data).

#### OTIS

The Optimized Complementary Feeding Study (OTIS) is a randomized controlled trial designed to evaluate the impact of a protein-reduced complementary diet based on Nordic foods. Recruitment started in April 2015 and ended in February 2019, including 250 infants. Measurements were gathered at approximately 5, 12, and 18 months of age. Anthropometric measurements, including weight and length, were recorded, and serum levels of insulin-like growth factor 1 (IGF1) were measured. In addition, saliva samples were obtained for genetic analyses. Further details can be consulted in the source publication of the study (55). The sample size for infants with genetic data is 200 individuals.

#### HOLBAEK

Formerly known as the Danish Childhood Obesity Biobank, the population-based cohort of the HOLBAEK Study included children and adolescents aged 4-20 years from schools across 11 municipalities in Zealand, Denmark. Individuals were enrolled between 2009 and 2019. The cohort is very well phenotyped, having data on anthropometrics, including height, weight, and BMI, as well as a broad range of metabolic biomarkers and protein measurements. Further details of the cohort have been published elsewhere (56,57). For this study, we used up to 632 samples with both proteomic and genetic data from children ranging 4 to 9 years of age.

### Phenotype measurement, processing, and standardization in MoBa

Height values at birth and pregnancy duration were obtained from hospital records through the Norwegian Medical Birth Registry (NMBR). Children’s length and height were measured at birth in the hospitals, and during routine visits by nurses at 6 weeks, 3, 6 and 8 months, and at 1, 1.5, 2, 3, 5, 7 and 8 years. Later, parents reported these measurements via questionnaires. This study used the version 12 of the curated questionnaires provided by MoBa. Extreme outliers defined as |value| > 5 × sd, e.g. due to handwritten errors or incorrect units, were excluded. Length/height values decreasing over time were considered aberrant and also excluded. Missing values preceded and followed by at least two values were imputed along the growth curve of children as done in Helgeland et al. (21). Height values were then standardized using the generalized additive model for location, scale and shape (GAMLSS) v5.4_22 (gamlss.com) in R 4.3.2 managed through conda environments in a similar way as in Helgeland et al. (21). Briefly, models based on a normal distribution were fitted for boys and girls separately using pregnancy duration as covariate for all time points. Models for ages until six months included a non-linear term for the pregnancy duration that was removed for later time points to ensure the convergence of GAMLSS models. Models were fitted on genetically unrelated children, defined as an identification by descent (IBD) coefficient lower than a third degree relative as inferred from the relatedness matrix provided with the MoBa genotypes. Z-scores were then computed for all children using the ‘centiles.pred’ function of GAMLSS.

### Genotyping and imputation

In MoBa, blood samples from the parents and their children (umbilical cord) were gathered at birth. The DNA was extracted and stored at the Norwegian Institute of Public Health. Samples have been genotyped in different batches through different research projects. The research projects that provided genetic data are HARVEST, SELECTIONpreDISPOSED, and NORMENT. The genotyping was done with a combination of different illumina microarrays. This study used quality controlled and imputed genotypes from the March 2023 MoBaPsychGen release (58). Specific details about the quality control for each batch are described in the relevant publication (58).

Briefly, of the total 238,001 individuals selected for genotyping, 235,412 were genotyped successfully and 234,505 samples of participants who maintained their consent were kept. The European Genome-Phenome Archive (Study ID EGAS00001001710) HRC release 1.1 was used for phasing and imputation. Whole chromosome phasing was done with SHAPEIT2, and imputation was done with IMPUTE4.1.2_r300.3. Post imputation, SNPs with INFO ≤ 0.8, MAF < 0.5, call rate 95%, HWE p < 1.00×10^-6^, and batch association with p-value < 5×10^-8^ were discarded. Samples were excluded if call rate < 98% and had ±3 standard deviations from the mean heterozygosity across all samples. After filtering, 76,577 children remained and 6,981,748 SNPs were available. A more detailed description about the specific QC steps and parameters is available in the relevant publication (58). The Genome Reference Consortium Human Build 37 (GRCh37) was used for all annotations.

Details regarding the genotyping of OTIS and HOLBAEK cohorts can be consulted in the supplementary material in the supplementary methods section.

### GWAS in MoBa

GWAS on standardized height values was performed at each time point using REGENIE v3.2.9 (59). Sex, pregnancy duration, plural births, and genotyping batch were used as covariates. A SNP was considered significant at any given time point if it passed a standard GWAS p-value threshold of 5×10^-8^. At each genome-wide significant locus and each time point, independent signals were obtained using conditional and joint (COJO) analysis as implemented in GCTA v1.93.2beta (41), yielding 548 SNPs in total. Since the same trait is evaluated across multiple time points, different SNPs identified at different time points might represent the same signal. To address this, pairwise colocalization analysis (61) was applied to all independent signals within a distance of ±500 kb across all time points using the R package COLOC v5.2.3. A graph was built using SNPs selected by COJO as vertices and COJO probabilities as edges. Two SNPs were considered to colocalize if p4 ≥ 0.8. Two SNPs were then considered independent if they did not colocalize and could not be connected by colocalizing SNPs. Among colocalizing SNPs, the one with lowest p-value was considered the lead variant, making 179 independent lead variants.

As secondary analyses, we conducted sex-stratified GWAS by dividing the sample into two subsets (girls and boys), using the same covariates as in the primary analysis, except for sex. Given the substantially increased multiple testing burden introduced by stratifying the data and analyzing each sex separately, we applied the Nyholt method (62) to estimate the effective number of independent tests based on phenotype correlations across the twelve timepoints (effective number = 4.62), yielding a corrected threshold of of p = 1.08×10^-8^ (5×10^-8^/4.62). Furthermore, since we tested three models (all individuals, male-only, and female-only), we applied an additional correction, resulting in a final genome-wide significance threshold of p = 3.6×10^-9^ (1.08×10^-8^/3).

Lastly, we also performed a genome-wide sex-interaction analysis, adjusting for the same covariates used in the primary discovery analysis, for which the standard GWAS significance threshold was applied. In addition, we examined whether any of the 179 independent SNPs identified in the main GWAS showed evidence of sex-interaction effects, using a Bonferroni-corrected threshold of p = 2.8×10^-4^(0.05/179).

### Clustering

To assess whether the effect sizes of the independent SNPs identified through COLOC v5.2.3 exhibited a temporal pattern, we performed hierarchical clustering using the *hclust* function in R 4.3.2, applied to the matrix of effect size estimates across time points. First, we extracted the effect sizes of the 179 independent variants from each GWAS performed in MoBa (n = 12). We also extracted the effect sizes for these SNPs from two GWASs performed in the UK Biobank: comparative height at age 10 (HAC10; n = 320,000, ukb-a-35) obtained from the IEU OpenGWAS project (63), and adult height (n = 480,000) previously published (44). The inclusion of these two UKBB GWAS in the clustering analysis was done to evaluate if there were variants whose effects weaken, prevail, or increase after eight years of age. Of note, the summary statistics from Yengo et al. (4) were not used due to the limited number of SNPs available (∼1 million). The alleles were aligned to the height-increasing allele at the age of peak association (lowest p-value) in MoBa GWAS.

Since the sample size was not consistent across GWAS, we transformed the effect sizes to sample-size corrected z-scores, as implemented by Suzuki et al. (65), with the following formula 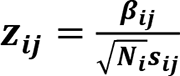, where ***s_ij_*** represents the standard error of the effect size estimate ***β_ij_*** for the phenotype i and SNP j, and ***N_i_*** is the corresponding sample size. Correlation was used as a dissimilarity metric calculated as follows: (**1** − ***ρ***)/**2**, where ρ is the correlation between two SNPs z-scores across all GWAS. The complete linkage algorithm was used to evaluate the distance between clusters. The optimal number of clusters was determined with the elbow method, resulting in the selection of three clusters. Lastly, we calculated the z-score average of all SNPs within each cluster for each time point to derive cluster-representative trajectories.

### LD score regression (LDSC) analyses

LDSC v1.0.1 (66) was used to calculate the SNP-based heritability (h2) using the pre-made LD scores of the European-ancestry 1000 Genomes project v3 reference. To calculate the genetic correlations within MoBa (n = 12) and other GWAS (n = 22), we used the cross-trait function implemented in LDSC (67). We used publicly available European GWAS summary statistics from the literature on anthropometric, metabolic, and puberty related traits: T2D (68), fasting glucose (69), fasting insulin (69), birthweight (36), adult BMI (64), adult height (64), childhood height at 10-12 years (5), Menarche (64). We also used UK Biobank GWAS summary statistics extracted from the IEU OpenGWAS project (63): Comparative height size at 10 years old (ukb-a-35), Body fat percentage (ukb-b-8909), Trunk fat percentage (ukb-b-16407), and relative age of first facial hair (ukb-a-299).

### SNP gene annotation and gene enrichment analyses

We annotated the SNPs to their closest gene with the R package snpsettest v0.1.2 (<u>github.com/HimesGroup/snpsettest</u>). The annotation was made with the gene curated GRCh37 reference available within the package. This reference has gene information from the GENECODE release 19, and only contains genes with known status and the following biotypes: protein-coding, Immunoglobulin (Ig), and variable chain and T-cell receptor (TcR) genes.

The genes assigned to the SNPs were subjected to gene enrichment analysis with the Web-based Gene SeT AnaLysis Toolkit (WebGestalt) 2019 (70). This online software has available different databases to perform enrichment analyses. We focused on disease related enrichment terms, of which they were curated with the Gene List Automatically Derived For You (GLAD4U) software (71). We evaluated enrichment with an overrepresentation analysis (hypergeometric-test), and accepted a false discovery rate (FDR) of 5%.

We also conducted a customized enrichment analysis by manually compiling gene panels of diseases potentially related to height, including musculoskeletal, growth, and glucose-related disorders using the UK gene panels based on rare variants (72) (panelapp.genomicsengland.co.uk). Here again a hypergeometric test and 5% FDR were used. We used R v4.3.2 for this analysis.

### Functional analysis (FUMA)

The SNPs mapped to nearest genes were subjected for enrichment analysis using the web based tool FUMA (Functional Mapping and Annotation of Genome-Wide Association Studies) (73). We used all sets of the background genes (protein coding, lncRNA, ncRNAm processed transcripts) corresponding to the GTEx v8 dataset (74). The correction for multiple testing was set at FDR < 5% and a minimum of 2 overlapped genes with the analyzed set.

### Proportion of the variance explained (PGS) in ALSPAC and MoBa

The potential variance explained by the identified SNPs in MoBa was tested in the ALSPAC cohort. Height measurements were binned based on the age of children resulting in 12 age bins 6 weeks, 9 months to 2 years, 3.5, 7, 8, 9, 10, 11, 12, 13, 15, and 17 years (supplementary table 1). In each bin, height was standardized to sex-specific z-scores by centering and scaling values using the mean and standard deviation, respectively. To test the variance explained by genetic variants, we first constructed polygenic risk scores (PGS) based on the SNPs of each cluster, and a PGS that contained all 179 independent SNPs. To construct the PGS, we used the effect sizes as weights, multiplied by the number of effect alleles, and summed over the SNPs. Then, we used linear models to calculate the variance explained (R2) of each PGS constructed at each time point. Both PGS calculations and linear regressions were done in R v4.3.2.

We also constructed and tested a PGS using the ∼10k of the ∼12k SNPs identified by Yengo et al. (4) that could be mapped in ALSPAC and MoBa. For this, we used the summary statistics from the European subset analysis, which included ∼1.5 million of individuals. The PGS was constructed with the score function implemented in PLINK2 (75). To further test how much of the variance could be explained by the adult-based PGS and parents height, we applied the following linear models across all the time points in MoBa: **Father height model**: Stature ∼ Father stature + sex; **Mother height model**; Stature ∼ Mother stature + sex; **Parents height model**: Stature ∼ Mother stature + Father stature + sex; **PGS model**: Stature ∼ PGS + sex and **PGS + parents height model**: Stature ∼ PGS + Mother stature + Father stature + sex.

### Determination of previously not reported variants for height

Variants were considered independent of known adult height or birth length hits if: (i) their p-value was strictly higher than 0.05/179 in the UKBB HAC10, UKBB adult height, and birth length (7) GWASs; and (ii) they were not in LD (R2 < 0.6) with any of the 12k SNPs reported to be associated with the largest adult height meta-analysis. The threshold 0.6 was chosen to exclude only hits in strong LD. The tool SNPclip in LDlink (76) was used for the LD analysis, using the aggregated European reference panels from the 1000 Genomes Project. Lastly, we performed a sign test to assess whether the potential novel loci showed consistent effect directions with those observed in ALSPAC. For this, we applied standard linear regression for each SNP, adjusting for sex. The analysis was conducted at age nine months, which best captures signals from the infancy growth cluster. We then evaluated whether the observed directional agreement was statistically significant using a binomial test.

We performed a manual database search for the genes annotated to the 19 novel loci to investigate their biological function. We investigated NCBI, OMIM, and UniProt by searching each gene individually and reviewing the summary pages. GWAS catalog was used to mine phenotypes related to height, height proxies and applicable cardio-metabolic traits. We used a reported p-value threshold of 5×10^-8^ for inclusion.

### Mendelian randomization analyses

To evaluate if there is a time-dependent causal effect of height with glucose related outcomes, we used both univariable and multivariable Mendelian randomization (MR). We assessed as exposures stature at birth, ages one, and seven from the MoBa cohort, as well as height at age 10 and in adulthood from the UK Biobank. The outcomes were T2D (68), fasting insulin (49), and fasting glucose (69). For the univariable MR we used the inverse variance weighted method (IVW) as the main analysis, while the MR EGGER and weighted median were used for sensitivity analyses. The instruments were the independent variants associated with height: in MoBa, independent SNPs were obtained from COJO, whereas for height at age 10 and adult height, SNPs were selected through clumping as implemented in PLINK2 (75). The clumping parameters were R2 < 0.001 within a window of ±500kb using the European 1000 Genomes Project as reference.

For the multivariable analysis MR (MVMR) we tested length at birth and height at one, seven and 10 years as exposures while adjusting for adult height in each model. We considered the MVMR IVW as the main analysis, and used both MVMR EGGER and MVMR weighted median for sensitivity analysis. For each time point, the instruments consisted of independent SNPs associated with height at that specific age, combined with independent SNPs associated with adult height. To avoid redundancy when merging these sets of SNPs, we applied clumping as described above. For both the univariable and multivariable analyses we used the R package "MendelianRandomization" v0.10.0 (77).

### Associations between of PGSs in ALSPAC, OTIS, and HOLBAEK

We evaluated the associations between the childhood-based (n=179 SNPs) and adult-based PGSs with metabolites and proteins relevant in cardiometabolic health available in ALSPAC, measured at childhood. The selected traits were: Fasting glucose (7 year), Fasting insulin (7 years), IGF1 (5 to 7 years), IGF2 (5 to 7 years), IGFBP3 (7 years), Leptin (8 years), GHBP (7 yeast) and SHGB (7 years). All measurements were obtained from blood samples. Details on the measurement procedures can be consulted in the ALSPAC variable dictionary, while a summary table of the corresponding study and the measurement units is available in the supplementary methods. We fitted linear models between the childhood or adult-based PGS and these traits, adjusted for sex, and we reported the variance explained (r2) corresponding to the PGS. Associations that passed FDR<0.05 were taken forward for replication in OTIS and HOLBAEK using the same modelling approach. IGF- related measurements in OTIS have been described elsewhere (55), and an overview of the variables available for HOLBAEK can be consulted in the supplementary methods. Sensitivity analyses were conducted across partitioned childhood clusters (infancy, childhood, lifetime, and combined infancy and childhood) to assess overall trends (p < 0.05) and identify which clusters may be driving the associations.

Since HOLBAEK also includes measurements of multiple metabolic health–related traits, and proteins, such as BMI, waist-to-hip ratio, and fasting glucose, we additionally conducted analyses across these relevant phenotypes. Full details are provided in the Supplementary Methods. Statistical significance was defined as FDR < 0.05, and sensitivity analyses were also performed by repeating these associations within the partitioned childhood clusters as performed in ALSPAC.

### Associations with the plasma proteome in HOLBAEK

We performed a hypothesis-free analysis of associations between childhood and adult PRS and plasma proteins measured in the HOLBAEK cohort (4–9 years), using Olink (149 proteins, n_children_= 633) and mass spectrometry (1,216 proteins, n_children_= 233). Associations between childhood height itself and plasma proteins were also assessed. Methodological details are described in the Supplementary Methods.

### Ethics

MoBa: Informed consent was obtained from all study participants. The administrative board of the Norwegian Mother, Father and Child Cohort Study led by the Norwegian Institute of Public Health approved the study protocol. The establishment of MoBa and initial data collection was based on a license from the Norwegian Data Protection Agency and approval from The Regional Committee for Medical Research Ethics. The MoBa cohort is currently regulated by the Norwegian Health Registry Act. The study was approved by The Regional Committee for Medical Research Ethics (#2012/67).

ALSPAC: Ethical approval for this study was obtained from the ALSPAC Ethics and Law Committee and the Local Research Ethics Committees. Study participants gave their informed consent for the use of the data obtained via questionnaires and clinics, following the recommendations of the ALSPAC Ethics and Law Committee. Consent for biological samples has been collected in accordance with the Human Tissue Act (2004).

HOLBAEK: The study was approved by the Ethics Committee of Region Zealand, Denmark (SJ-104) and by the Danish Data Protection Agency (REG-043-2013). The HOLBAEK Study is registered at ClinicalTrials.gov (NCT00928473). Written informed consent was obtained from all participants in accordance with the Declaration of Helsinki. For participants younger than 18 years, informed oral assent was obtained from the participant and written informed consent was provided by their parents or legal guardians.

OTIS: The study was approved by the Regional Ethical Review Board at Umeå University (2014-363-31M and 2018-4-32), Umeå, Sweden and is registered at <u>ClinicalTrials.gov</u> (NCT02634749). Written informed consent was obtained from both caregivers.

## Acknowledgements

This work was supported by grants to S.J from Helse Vest’s Open Research Grant (#912250 and F-12144), the Novo Nordisk Foundation (NNF20OC0063872) and the Research Council of Norway (#315599) and by the Meltzer Research Fund.

K.K.O. was supported by the UK Medical Research Council (MC_UU_00006/2) and the NIHR Cambridge Biomedical Research Centre. M.V. was supported by the Research Council of Norway (#301178), the European Research Council (#101171420), and the University of Bergen.

GDS works within the MRC Integrative Epidemiology Unit at the University of Bristol, which is supported by the Medical Research Council (MC_UU_00032/1).

MoBa: We thank the Norwegian Institute of Public Health (NIPH) for generating the high quality genomic data. This research is part of the HARVEST collaboration, supported by the Research Council of Norway (#229624). We also thank the NORMENT Centre for providing genotype data, funded by the Research Council of Norway (#223273), South East Norway Health Authorities and Stiftelsen Kristian Gerhard Jebsen, and in collaboration with deCODE Genetics. We further thank the Center for Diabetes Research, the University of Bergen for providing genotype data and performing quality control and imputation of the data funded by the ERC AdG project SELECTionPREDISPOSED, Stiftelsen Kristian Gerhard Jebsen, Trond Mohn Foundation, the Research Council of Norway, the Novo Nordisk Foundation, the University of Bergen, and the Western Norway Health Authorities. We thank the MoBaPsychGen team, led by Elizabeth Corfield, for providing quality controlled genotype data, supported by funding from the South-Eastern Norway Regional Health Authority (#2021045; #2020022; #2022083; 2018058).

We are grateful to all the families in Norway who are taking part in the ongoing MoBa cohort study. All analyses were performed using digital labs in HUNT Cloud at the Norwegian University of Science and Technology, Trondheim, Norway. We are grateful for outstanding support from the HUNT Cloud community.

ALSPAC: We are extremely grateful to all the families who took part in the ALSPAC cohort study, the midwives for their help in recruiting them, and the whole ALSPAC team, which includes interviewers, computer and laboratory technicians, clerical workers, research scientists, volunteers, managers, receptionists and nurses. The UK Medical Research Council and Wellcome (217065/Z/19/Z) and the University of Bristol provide core support for ALSPAC. This publication is the work of the authors and S.J. will serve as guarantor for the contents of this paper. A comprehensive list of funding is available on the ALSPAC website (www.bristol.ac.uk/alspac/external/documents/grant-acknowledgements.pdf); this research was specifically funded by Wellcome Trust and MRC (core) (076467/Z/05/Z). ALSPAC GWAS data was generated by Sample Logistics and Genotyping Facilities at Wellcome Sanger Institute and LabCorp (Laboratory Corporation of America) using support from 23andMe.

P.S.N received funding from The Swedish Research Council, Stockholm, Sweden (2023–02735), The Swedish Society for Medical Research (SG-24-0105-B) and the Agreement concerning research and education of doctors (ALFGBG-1005149). B.J. received funding from The Swedish Research Council, Stockholm, Sweden (2019-01004 and 2024-02502), The Research Council of Norway (RCN), Oslo, Norway (FRIMEDBIO #547711), March of Dimes (#21-FY16-121), Agreement concerning research and education of doctors (ALFGBG-965353 and ALFGBG-1005151). OAA received funding from RCN (#324499, #326813) and Nordforsk (#164218).

Y.H and L.A.H. received research grants from the Danish Cardiovascular Academy, which is funded by the Novo Nordisk Foundation (grant no. NNF20SA0067242) and the Danish Heart Foundation. The HOLBAEK study was supported by the Innovation Fund Denmark (060300484B), the Novo Nordisk Foundation (NNF15OC0016544).

## Contributions

SJ and NFB designed and conceptualized the study. AEL contributed to the study design. NFB, SJ, AEL and MV wrote the manuscript. NFB and AEL performed the core statistical analyses. TH, LAH, JCH designed the HOLBAEK sample. ICP and YU curated the data and performed statistical analyses in HOLBAEK. JS and RK performed data analyses and interpretation. TL designed the OTIS cohort, and NFB performed the statistical analyses. AH and OAA conceptualized and curated the MoBa PsychGen release. SO’R, PSN, BJ, PJ, KKO, DSG, EB, MV and NPR contributed with data and/or analysis and interpretation. SJ and MV supervised the work. All the authors critically reviewed the manuscript.

## Data availability

Data from the Norwegian Mother, Father and Child Cohort Study used in this study are managed by the Norwegian Institute of public health and can be made available to researchers, provided approval from the Regional Committees for Medical and Health Research Ethics (REC), compliance with the EU General Data Protection Regulation (GDPR) and approval from the data owners. The consent given by the participants does not open for storage of data on an individual level in repositories or journals. Researchers who want access to data sets for replication should apply through helsedata.no. Access to data sets requires approval from The Regional Committee for Medical and Health Research Ethics in Norway and an agreement with MoBa.

The informed consent obtained from ALSPAC (Avon Longitudinal Study of Parents and Children) participants does not allow the data to be made available through any third party maintained public repository. Supporting data are available from ALSPAC on request under the approved proposal number, BB3605. Full instructions for applying for data access can be found here: www.bristol.ac.uk/alspac/researchers/access. The ALSPAC study website contains details of all available data (www.bristol.ac.uk/alspac/researchers/our-data).

In accordance with the General Data Protection Regulation (GDPR, https://gdpr-info.eu/), individual-level clinical and proteomics data in the HOLBAEK study cannot be made publicly available to maintain patient confidentiality, but are available upon request to T.H. at, and J.C.H. at. Access to the data can be granted through the Danish Data Protection Agency and the ethics committee for the Region Zealand, Denmark, subject to obtaining proper approvals and adherence to patient information and data-processing agreements.

OTIS data is available upon reasonable request by contacting the corresponding authors and T.L.

