## Supplementary material for "The Genetic and Proteomic Determinants of Pediatric Stature Development and their link to adult height and Type 2 Diabetes"

### Supplementary methods

### ALSPAC supplementary methods

#### *Genotyping*

ALSPAC genotyping has been thoroughly described elsewhere and details can be consulted either in its source publication (1) or their official website (https://proposals.epi.bristol.ac.uk/alspac_omics_data_catalogue.html)

#### *Metabolite and protein data from*

The ALSPAC cohort offers data on several phenotypic measures, for which detailed information can be found in the variable dictionary (https://variables.alspac.bris.ac.uk/). Table 1 provides an overview of the variables analyzed in relation to child- and adult- based polygenic scores (PGS). All data was measured from blood.

Table 1: Information of ALSPAC variables used for this study.

| Dataset | Variable | Details | Data source | Generation |
|---|---|---|---|---|
| **Child_bloods_7a** | Glucose_BBS | Fasting glucose mmol/l, BBS | biosamples | G1 |
| **Child_bloods_7a** | Insulin_BBS | Fasting insulin mu/l, BBS | biosamples | G1 |
| **Child_bloods_7a** | IGF1_CIF61 | IGF I ng/ml, 61mth CIF | biosamples | G1 |
| **Child_bloods_7a** | IGF1_F7 | IGF1, F7 | biosamples | G1 |
| **Child_bloods_7a** | IGF2_F7 | IGF2, F7 | biosamples | G1 |
| **Child_bloods_7a** | IGF2_CIF61 | IGF II ng/ml, 61mth CIF | biosamples | G1 |
| **Child_bloods_7a** | IGFBP3_F7 | IGFBP3, F7 | biosamples | G1 |
| **Child_bloods_7a** | LEPTIN_f9 | Leptin ng/ml, Focus 9 years | biosamples | G1 |
| **Child_bloods_7a** | GHBP_BBS | Growth hormone binding protein ng/ml, BBS | biosamples | G1 |
| **Child_bloods_7a** | SHBG_BBS | Sex hormone-binding globulin nmol/l, BBS | biosamples | G1 |

G1 refers to the children of the G0 mothers. CIF61 indicates that data were collected at 61 months of age (approximately 5 years). F7 and F9 denote assessments conducted at ages 7 and 9 years, respectively. BBS refers to the Before Breakfast Study, which collected biosamples from children at 7 years of age. The data was extracted from the ALSPAC variable search tool website (https://variables.alspac.bris.ac.uk/).

### OTIS supplementary methods

#### *Genotyping*

Genotyping was done with the Illumina HumanCoreExome platform. Genotypes were called in Illumina GenomeStudio (v2011.1). Cluster positions were identified from samples with call rate ≥0.98 and GenCall score ≥ 0.15. We excluded variants with call rate < 98%, cluster separation < 0.4, 10% GC score < 0.3, AA T Dev > 0.025, Hardy–Weinberg equilibrium P value < 1 × 10−6. Samples were, and heterozygosity excess of > 4 s.d. Values.

### HOLBAEK supplementary methods

#### *Sample size and variables*

This study used data from the population-based cohort of The HOLBÆK Study, including children and adolescents recruited from schools across the Zealand region, Denmark. All participants underwent deep cardiometabolic phenotyping, as described previously (2). Anthropometric measurements, including height, weight, waist circumference, and hip circumference, were obtained by trained medical professionals. Body mass index standard deviation scores (BMI SDS) were calculated according to Danish reference data, while waist-to-height ratio (WHtR) SDS and waist-to-hip ratio (WHR) SDS were derived using international age- and sex-specific reference standards. Systolic and diastolic blood pressure standard deviation scores (SBP SDS and DBP SDS) were derived using age-, sex-, and height-specific reference values. Venous blood samples were collected after an overnight fast. Biochemical measurements included plasma concentrations of alanine aminotransferase (ALT), aspartate aminotransferase (AST), alkaline phosphatase (ALP), gamma-glutamyl transferase (GGT), bilirubin, HDL cholesterol (HDL-C), LDL cholesterol (LDL-C), total cholesterol, triglycerides (TG), and glucose; serum concentrations of insulin and C-peptide; and whole-blood glycated hemoglobin (HbA1c). A total of 634 individuals aged 2–10 years from the population-based cohort were included. The mean age was 7.96 ± 1.21 years, and 52.5% were female.

#### Proteomic profiling

Plasma proteomic profiling was performed using two complementary platforms: Olink proximity extension assays including Target 96 Cardiovascular II (CVDII) and Target 96 Inflammation (INF) panels, and mass spectrometry (MS)-based platform. Proteins measured using Olink were reported as normalized protein expression (NPX) values on a log2 scale. A total of 149 proteins detected above the limit of detection in more than 80% of individuals were included. The MS-based proteomics dataset included 1,216 proteins with 91% completeness. Protein abundances were $\log_2$ -transformed and corrected for sample preparation batches prior to analysis. Details of preprocessing and quality control have been described previously (2,3).

**Genotyping, PGS characteristics and construction**

Genotyping was performed using Illumina Infinium HumanCoreExome BeadChips, followed by standard quality control, phasing, and imputation using the Sanger Imputation Server with the HRC1.1 reference panel. Individuals and variants failing quality control (e.g., low call rate, low imputation quality, or non-European ancestry) were excluded. Details have been described previously (3).

Six polygenic risk scores (PGS) were constructed: PGSs based on each of the three SNP clusters (infancy, childhood, and lifetime), and a PGS including all 179 independent SNPs. In addition, an adult PGS based on 3,290 variants (4), and a composite PGS combining the infancy and childhood components were generated. All PGSs were calculated as weighted sums of risk alleles and standardised prior to analysis.

**Statistical analyses**

All statistical analyses were performed in R. Cardiometabolic traits with right-skewed distributions were log-transformed, and subsequently standardised (z-scored) prior to modelling to obtain comparable effect estimates across traits.

Associations between PGS and cardiometabolic traits were assessed using linear regression models. Effect estimates are reported as standardised β coefficients, representing the change in outcome (in SD units) per 1 SD increase in PGS. All models were adjusted for age, sex, genotyping batch, and the first four genetic principal components (PC1–PC4):

OUTCOME ~ PGS + age + sex + batch + PC1 + PC2 + PC3 + PC4

Analyses were restricted to individuals aged 2–10 years and performed within the population-based cohort.

Associations between PGS and circulating protein levels were analysed using the same modelling framework.

PROTEIN ~ PGS + age + sex + batch + PC1 + PC2 + PC3 + PC4

Finally, two additional sets of models were fitted to examine associations between height SDS and circulating proteins. These models included age and sex as covariates, with additional adjustment for waist-to-hip ratio SDS:

PROTEIN ~ HEIGHT SDS + age + sex (+ WHR SDS)

**REFERENCES SUPPLEMENTARY METHODS**

1. Horikoshi M, Yaghootkar H, Mook-Kanamori DO, Sovio U, Taal HR, Hennig BJ, et al. New loci associated with birth weight identify genetic links between intrauterine growth and adult height and metabolism. Nat Genet. 2013 Jan 1;45(1):76–82. doi:10.1038/ng.2477

2. Stinson SE, Huang Y, Thielemann R, Stankevic E, Lund MAV, Holm LA, et al. Identification of modifiable plasma protein markers of cardiometabolic risk in children and adolescents with obesity. Nat Commun. 2026 Jan 14;17(1):1718. doi:10.1038/s41467-026-68415-2

3. Niu L, Stinson SE, Holm LA, Lund MAV, Fonvig CE, Cobuccio L, et al. Plasma proteome variation and its genetic determinants in children and adolescents. Nat Genet. 2025 Mar 1;57(3):635–46. doi:10.1038/s41588-025-02089-2

4. Yengo L, Sidorenko J, Kemper KE, Zheng Z, Wood AR, Weedon MN, et al. Meta-analysis of genome-wide association studies for height and body mass index in ∼700000 individuals of European ancestry. Hum Mol Genet. 2018 Oct 15;27(20):3641–9. doi:10.1093/hmg/ddy271

**Supplementary figures**

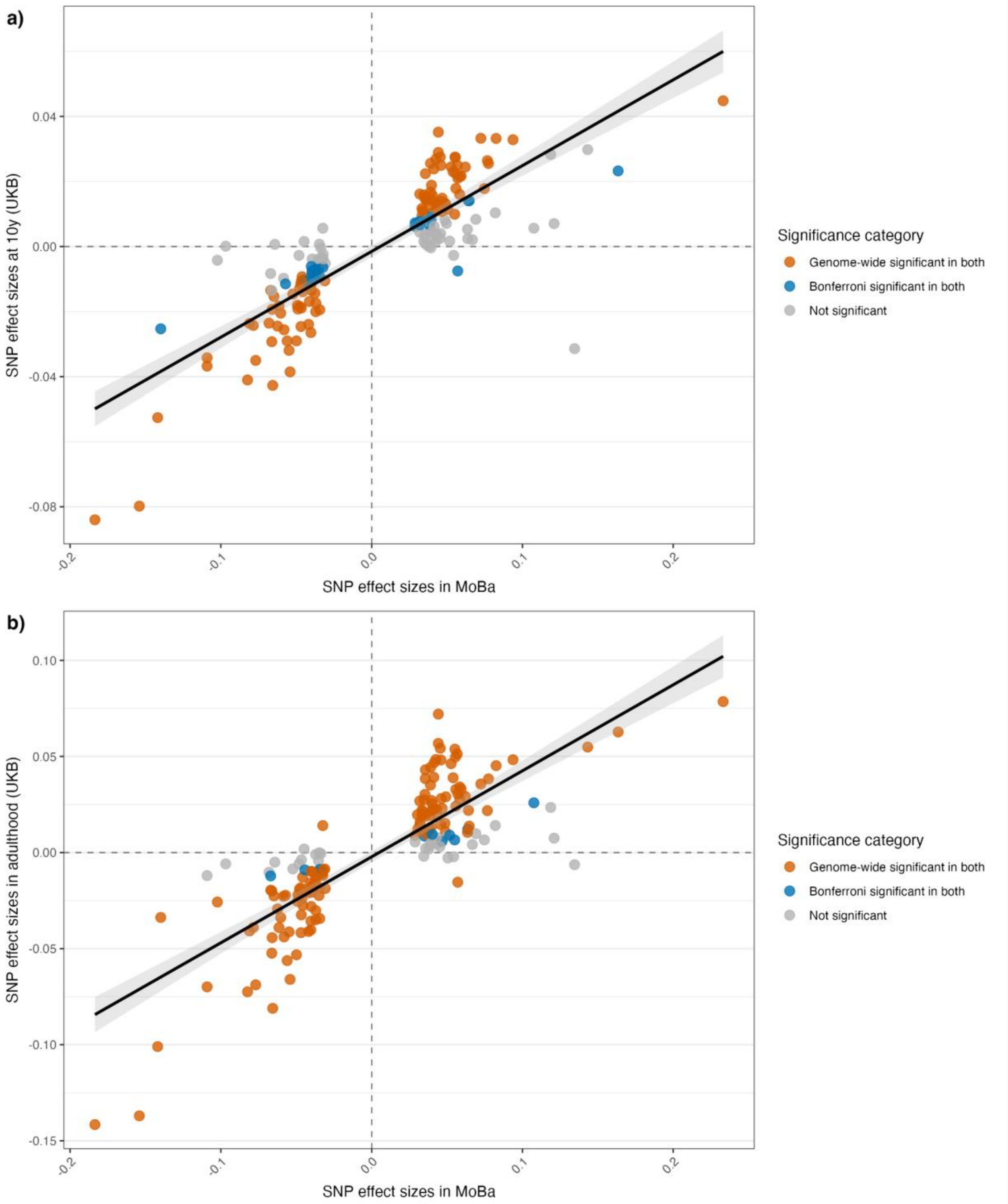


**Supplementary figure 1: Effect size comparison of the 179 SNPs with UKB GWAS:** Effect size at peak child-effect time point in MoBa compared with a) comparative height size at 10 years and b) adult height.

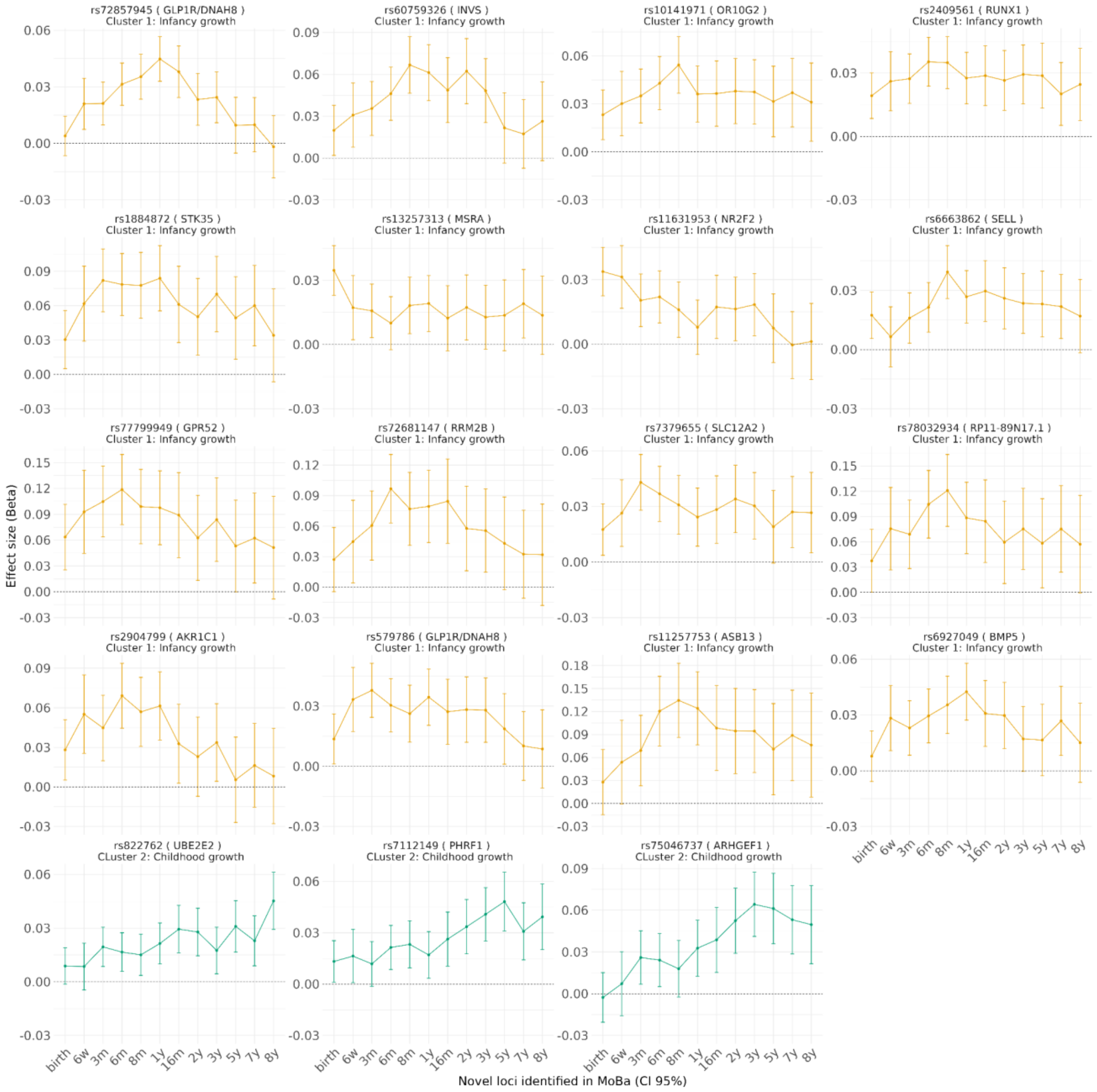


**Supplementary figure 2: Individual trajectories of the novel SNPs.** Effect trajectories are illustrated for a representative phenotype under the classical additive model (teal), as well as under the sex-stratified model—salmon for females and green for males—across time.

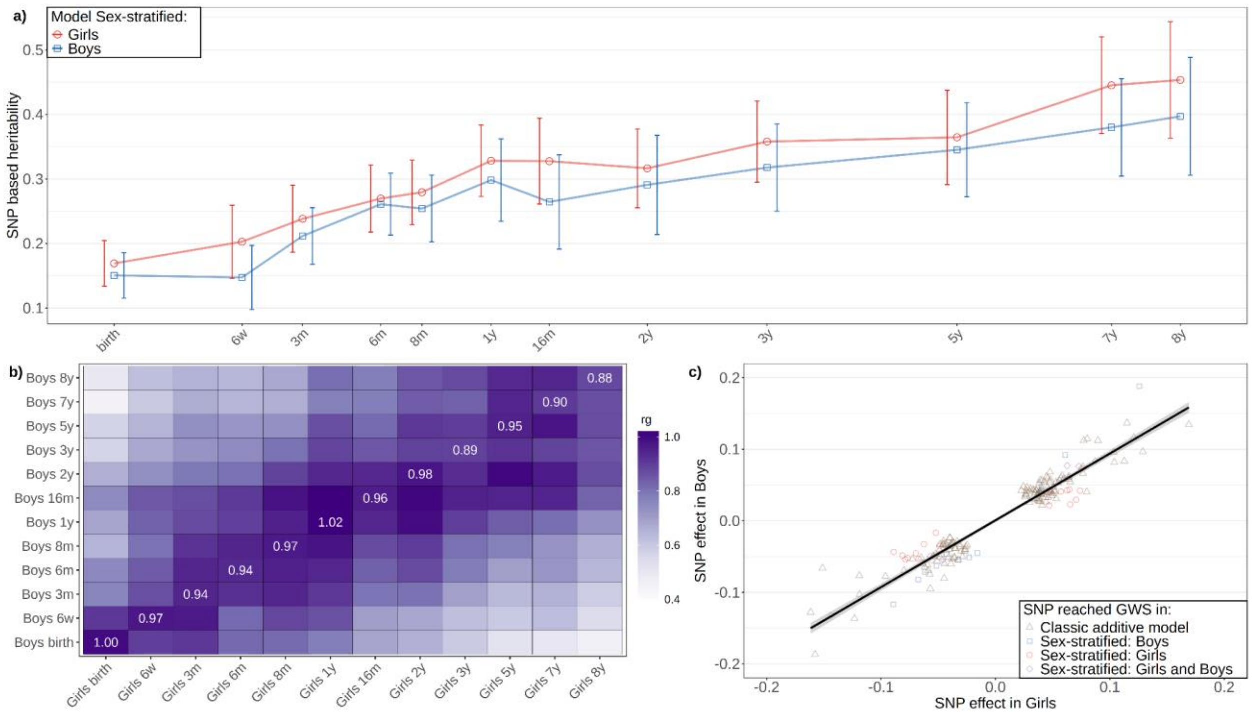


**Supplementary figure 3: Sex specific GWAS of childhood height.** a) SNP-based heritability in girls and boys (95% CI), b) genetic correlation between the sex-stratified analyses across time-points, c) comparison between girls and boys of sex-stratified effect estimates (additive models) of the 179 lead SNPs.

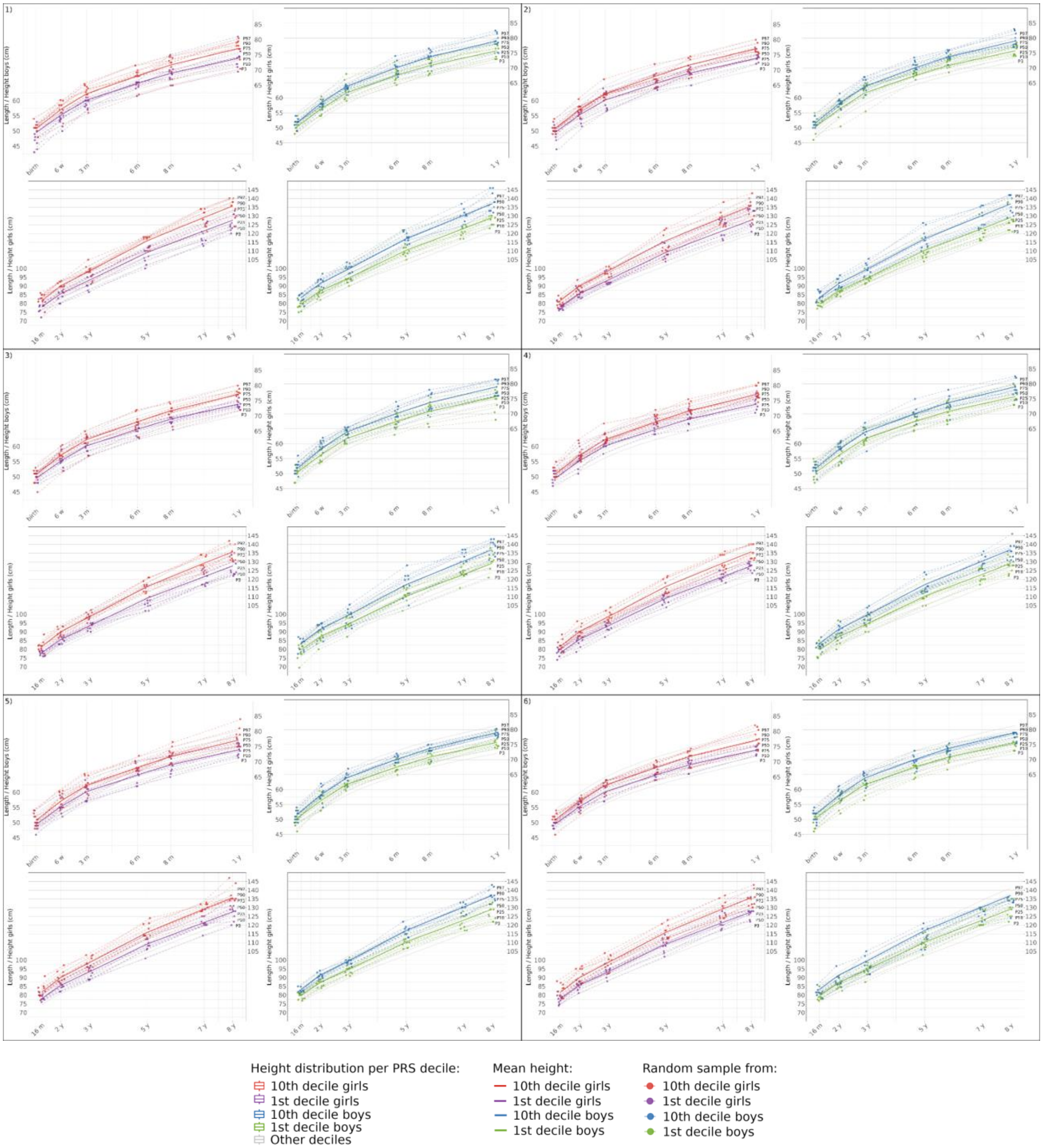


**Supplementary figure 4: Height distribution across PGS deciles and longitudinal trajectories of a random draw of individuals in the first and tenth deciles in the MoBa cohort.** Each iteration shows the longitudinal trajectories of a random draw of 10 individuals from the 1st and 10th deciles in the MoBa Cohort by sex. Height distribution by decile in girls. Individual trajectories (dashed lines) from birth to 1 year old of 5 girls from the first decile (purple) and 5 from the tenth (red). Individual trajectories (dashed lines) of girls from 16 months to 8 years old from the first decile (purple) and from the tenth (red). The solid lines indicate the mean of the deciles in their respective color, d) Height distribution by decile in boys. Individual trajectories (dashed lines) from birth to 1 year old of 5 boys from the first decile (green) and 5 from the tenth (blue). Individual trajectories (dashed lines) of a random draw of boys from 16 months to 8 years old from the first decile (green) and from the tenth (blue). The solid lines indicate the mean of the deciles in their respective color. See Supplementary figure 4 for other random sample draws to illustrate the consistent height growth pattern in individuals from 1st and 10th decile.

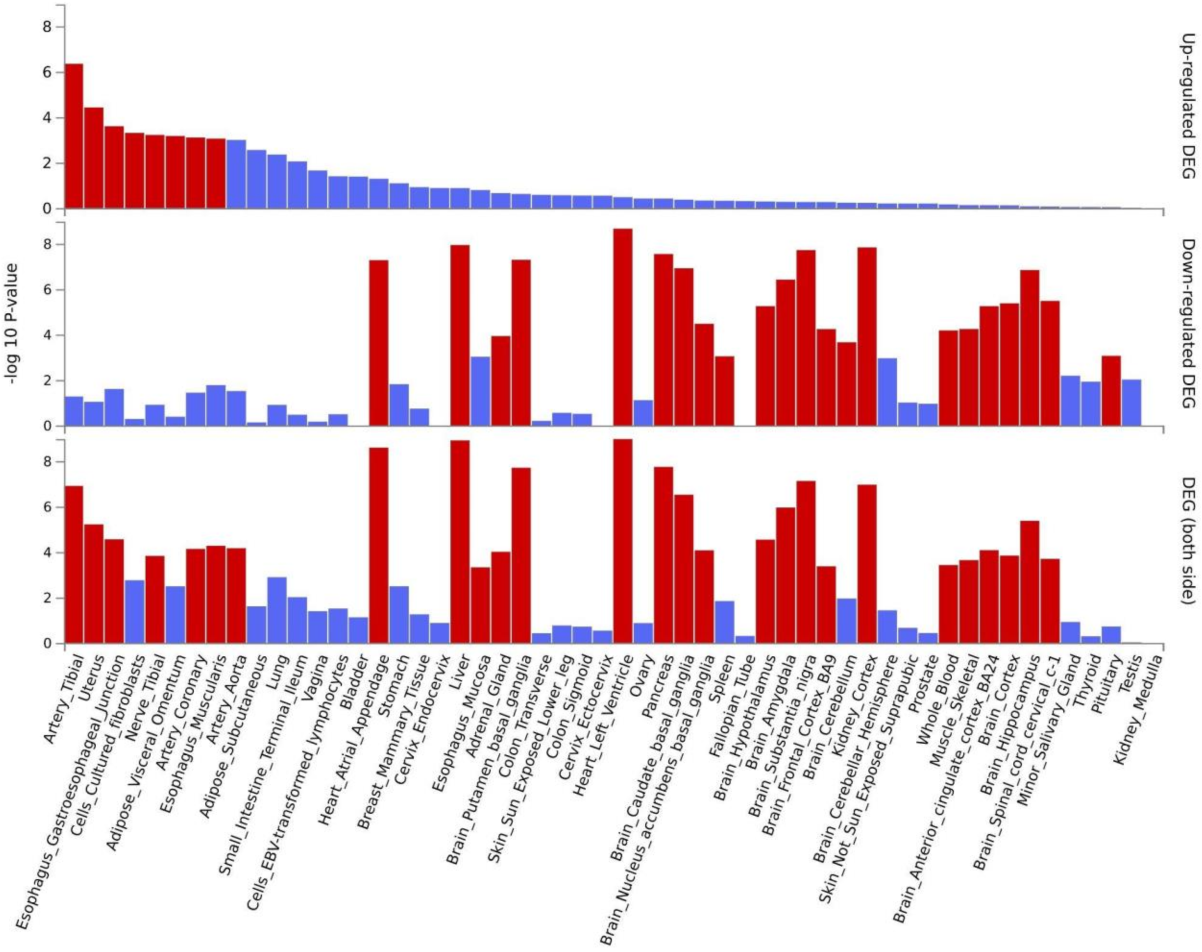

**Supplementary figure 5: Differentially expressed genes in 54 GTEx tissues.** Pre-calculated sets of differentially expressed genes (DEGs) from Genotype-Tissue Expression (GTEx) were used for each expression dataset. To define each DEG set, genes were identified using a two-sided t-test comparing the expression of each label with all other groups. A gene was considered differentially expressed if it had a Bonferroni-corrected p-value less than 0.05 and an absolute log fold change of at least 0.58. For signed DEG sets, genes were further classified by the direction of expression. The up-regulated DEG set includes genes significantly overexpressed in samples with that label compared to other samples. The -log10(P values) in the graph refer to the probability calculated from the hypergeometric test.
